# National PReCePT Programme: a quasi-experimental before-and-after evaluation of the implementation of a national quality improvement programme to increase the uptake of magnesium sulphate in pre-term deliveries

**DOI:** 10.1101/2022.05.20.22275353

**Authors:** Hannah B Edwards, Maria Theresa Redaniel, Carlos Sillero-Rejon, Ruta Margelyte, Tim J Peters, Kate Tilling, William Hollingworth, Hugh McLeod, Pippa Craggs, Elizabeth M Hill, Sabi Redwood, Jenny L Donovan, Emma Treloar, Ellie Wetz, Natasha Swinscoe, Gary A Ford, John Macleod, Karen Luyt

## Abstract

**Objective:** To evaluate the effectiveness and cost-effectiveness of the National PreCePT Programme (NPP) in increasing magnesium sulphate (MgSO_4_) in pre-term births.

**Design:** A quasi-experimental before-and-after design

**Setting:** Maternity units within NHS England and the AHSN network in 2018.

**Participants:** Maternity units in England (n=137) who participated in the NPP.

**Interventions:** NPP support which included the PReCePT QI toolkit and materials (pre-term labour proforma, staff training presentations, parent leaflet, posters for the unit, learning log), regional AHSN level support, and up to 90 hours funded backfill for a midwife ‘champion’ to lead implementation.

**Main outcome measures:** MgSO_4_ post-implementation uptake compared to pre-implementation uptake. Implementation and lifetime costs were estimated.

**Results:** Compared to pre-implementation estimates, the average MgSO_4_ uptake in 137 maternity units in England increased by 6.3 percentage points (95% CI 2.6 to 10.0 percentage points) to 83.1% post-implementation, accounting for unit size, maternal, baby, and maternity unit factors, time trends, and AHSN. Further adjustment for early or late initiation of NPP activities increased the estimate to 9.5 percentage points (95% CI 4.3 to 14.7 percentage points). From a societal and lifetime perspective, the health gains and cost-savings associated with the NPP effectiveness generated a net monetary benefit of £866 per preterm baby and the probability of the NPP being cost-effective was greater than 95%.

**Conclusion:** This national QI programme was effective and cost-effective. National programmes delivered via coordinated regional clinical networks can accelerate uptake of evidence-based therapies in perinatal care.

**WHAT IS ALREADY KNOWN ON THIS TOPIC:** Since 2015 the UK National Institute for Health and Care Excellence (NICE) has recommended administration of Magnesium Sulphate (MgSO_4_) for fetal neuroprotection in very preterm deliveries as a core part of maternity care. By 2017, only two-thirds of all eligible women in England were being given MgSO_4_, with wide regional variation. The PReCePT pilot study showed an increase in uptake from 21% (2012-2013) to 88% after the introduction of the tool in2015 in 5 units. The National PReCePT Programme (NPP) was rolled out in 2018 by the national network of Academic Health Science Networks with a target to increase MgSO_4_ administration in England to 85% by 2020.

**WHAT THIS STUDY ADDS:** The study has shown that the nationwide implementation of the NPP, which provided a Quality Improvement (QI) toolkit and materials, cross unit AHSN regional level support, and funded backfill for a unit midwife ‘champion’ through local regional support was effective and cost-effective. Scaling-up of network supported QI programmes at national level can accelerate uptake of new therapies and promote improvements in perinatal care delivery.

## INTRODUCTION

Around 10% of very preterm births in developed countries result in Cerebral Palsy (CP),^1-3^ representing a significant burden for affected individuals and families,^4^ and costs for healthcare services. Antenatal magnesium sulphate (MgSO_4_) reduces the risk of CP in the child by around 30% when given to women at risk of preterm birth,^5^ and around 200 cases of CP in England could be avoided by consistent administration of MgSO_4_ during labour each year.^6^ Administration of MgSO4 can be highly cost-effective with an estimated £1M lifetime societal savings per case of CP averted.^7,8^

Since 2015 the UK National Institute for Health and Care Excellence (NICE) has recommended administration of MgSO_4_ in very preterm deliveries as a core part of maternity care.^9^ Yet, by 2017 only 64% of eligible women were receiving MgSO_4_.^10^ High regional variation in uptake (range 49% to 78%) indicated inequalities in perinatal care.^10^

The PReCePT (Preventing Cerebral Palsy in Pre Term labour) Quality Improvement (QI) toolkit and approach was developed to improve maternity staff awareness and thereby increase use of MgSO_4_, and the pilot study (five maternity units) showed an increase in uptake from 21% to 88% after the completion of the pilot.^11^ In 2018, NHS England funded the National PReCePT Programme (NPP), with units receiving implementation support provided regionally through the 15 Academic Health Science Networks (AHSNs), with the aim of increasing MgSO_4_ use in England to 85% by 2020.

In the NPP, the PReCePT QI toolkit and other materials (pre-term labour proforma, staff training presentations, parent information leaflet, posters for the unit, and a learning log)^12^ were provided to each participating unit (see supplementatry file - National PReCePT Programme provisions). Each unit appointed a lead midwife to be their ‘PReCePT champion’, and the NPP provided up to 90 hours funded backfill for this role. AHSN level coaching and support via a clinical lead (obstetrician and/or neonatologist) was also available to each unit and champion. Additional support was provided by the AHSN Network NPP steering group and the national Clinical Lead and Programme Manager, and through shared learning events between AHSN managers leading the NPP in their area. The NPP was launched in two tranches of maternity units, in May and September 2018. A cluster randomised trial to determine the effectiveness of an enhanced support QI programme (ESP) was conducted alongside the NPP implementation comparing uptake in 13 ESP units and 27 control units implementing the NPP.^13,14^

In this study we evaluated the effectiveness and cost-effectiveness^15^ of the NPP in increasing MgSO_4_ uptake. We hypothesised that the implementation of the NPP would help increase MgSO_4_ uptake beyond the expected increase due to the underlying trend rate.

## METHODS

The evaluation followed a quasi-experimental before-and-after design, comparing absolute difference in mean MgSO_4_ uptake between the 12 months pre-implementation and 12 months post-implementation periods, adjusted for possible confounders. The month of the initiation of the NPP in the maternity unit (adoption date) was excluded from the analysis.

### Data

We used anonymised patient-level data from the UK National Neonatal Research Database (NNRD).^16^ The database collates information from electronic patient records of neonatal units who are part of the UK National Neonatal Collaborative. Data collected include mother and baby socio-demographic characteristics, clinical information and data on MgSO_4_ use. All data in the NNRD undergoes multiple quality assurance procedures and is considered to have high accuracy and completeness.^16,17^ Fields relating to the MgSO_4_ care pathway are regarded as good quality (over 70% completeness).

Estimated NPP adoption dates for each unit, demarcating the two periods, were provided by the AHSNs. This was defined as the date when the unit had initiated an implementation plan and patients started benefitting from the Programme. Pre- and post-implementation periods covered between October 2017 and June 2020.

### Outcome

In the effectiveness analysis, for consistency with nationally reported data (National Neonatal Audit Programme (NNAP) reporting), MgSO_4_ uptake was defined as the number of mothers recorded as receiving MgSO_4_ divided by the total number of eligible mothers, excluding missing values from the denominator. This was expressed as a percentage and computed per month per unit. For the cost-effectiveness analysis, missing MgSO_4_ uptake was considered as ‘not given’ and included in the denominator.

We only included data on singletons and one baby (the first born) from each multiple birth. All babies in multiple births with an NNRD record were included in descriptions of baby-level demographics. In cases where only one baby had a record for MgSO_4_, we recoded the missing MgSO_4_ status of the other multiples to match that for their twin/triplet who did have a record. For multiples with conflicting records (e.g. Baby1=given, Baby2=not given), we recorded MgSO_4_ uptake as having been given.

Secondary outcomes were the trends in MgSO_4_ uptake and missingness in MgSO_4_ data across the whole NPP period, (where available) reasons MgSO_4_ was recorded as not given, costs of the NPP per preterm baby and the incremental net monetary benefit of the NPP per preterm baby from a lifetime societal perspective.

### Possible confounders and other model terms

Possible confounding factors adjusted for in the study included variables at three levels: the baby, mother and maternity unit. Characteristics of the baby were: birthweight (grams) adjusted for gestational age (weeks) and sex, expressed as a z-score; and whether the baby was part of a multiple birth (yes/no). Maternal characteristics were: age at the time of the birth (years); ethnicity (white or black/asian/mixed); level of deprivation (Index of Multiple Deprivation (IMD) decile); and maternal hypertension during pregnancy (yes/no). The maternity unit level factor was type of unit (High Dependency or Special Care Unit (HDU/SCU) versus Neonatal Intensive Care Units (NICU)). Also taken into account, were type of birth (imminent or threatened, only for cost-effectiveness analysis), time (study month, included as a linear variable, with time represented as the cumulative number of months since 12-months pre-implementation), and clustering by AHSN. The possible confounders were selected based on the literature and on the recommendation of clinical experts.

For the effectiveness analysis, all baby and mother characteristics were aggregated to the level of the maternity unit (using non-missing information) per study month as follows: mean z-score for gestational age adjusted for birthweight, proportion of multiple births of all eligible births, mean maternal age, proportion of mothers who identify as of white ethnicity, mean IMD decile, proportion of mothers who had pregnancy hypertension. Missing information was minimal, with the exception of mother’s ethnicity (supplementary file - Missing data on possible confounders). The cost-effectiveness analysis used baby-level data, and missing data on possible confounders were imputed through chained equations.^18^

### Statistical analyses

#### Effectiveness analysis

To compare the difference in mean monthly MgSO_4_ uptake between the pre- and post-implementation periods, we conducted a multi-level mixed-effects linear regression using the maternity unit as the primary level of analysis. The mean MgSO_4_ uptake was modelled directly; the model was weighted on unit size (number of eligible mothers at each unit) and adjusted for clustering by AHSN (secondary level of analysis). The model was adjusted for all known potential confounders.

To take into account NPP activities prior to the reported NPP implementation dates (as units were reported to have started with some activities before or after the adoption month), weexcluded records within three months either side of the NPP adoption month.

As sensitivity analyses, we also looked at the effect of the following on the difference in mean monthly MgSO_4_ uptake between the two NPP periods: (1) including the 13 PReCePT ESP trial units that received the enhanced QI support programme (NPP plus unit-level coaching, an additional 90 hours backfill funding for the local midwife champion and approximately 104 hours of funded backfill for the unit obstetrician/neonatologist lead);^13^ and (2) excluding units in one AHSN that started adoption three months later than other AHSNs.

#### Cost-effectiveness analysis

The mean implementation cost per unit of the NPP was estimated from data supplied by the national programme team. NPP costs included those relating to national programme management, funded support from AHSNs, and the funded backfill of clinical time for midwives and regional clinical leads. The mean implementation cost per baby was calculated as the mean implementation cost per unit divided by the total number of babies per unit eligible for MgSO_4_ treatment delivered during the 12-month follow-up period.

In a decision tree analysis, we estimated the net monetary benefit of the NPP using a lifetime horizon and societal perspective. Model parameters were based on NNRD data for MgSO_4_ uptake, and reported estimates for lifetime gains in quality-adjusted life-years (QALYs) and societal cost savings as a consequence of preventing CP via MgSO_4_ treatment.^19^ All cost estimates were converted to Pounds Sterling and inflated to 2019 prices (supplementary file - Estimated lifetime costs and QALYs per patient associated with MgSO4 treatment). Babies delivered by caesarean section were defined as imminent births (those certain to occur within 24 hours) and all other babies as threatened. Our deterministic analysis used a willingness-to-pay threshold of £20,000 per QALY gained, in line with the relevant NICE guidance.^20^

To estimate the model parameter associated with MgSO_4_ uptake differences between the baseline and follow-up periods, we conducted a multi-level mixed-effects linear logistic regression at the baby level, adjusted for clustering by AHSN and unit. The model was also adjusted for all listed confounders, including type of birth and interaction between type of birth and pre- and post-implementation, entered into the model simultaneously.

We conducted a probabilistic analysis using Monte Carlo simulation with 10,000 samples drawn from the parameter distributions. For lifetime costs and health utilities estimates, we used the incremental differences. Point estimates, probabilistic distribution assumptions, and parameter source estimates are reported in appendix p 10. Incremental costs and effects were plotted on the cost-effectiveness plane and a cost-effectiveness acceptability curve plotted for willingness-to-pay thresholds from zero to £100,000 per QALY gained. In a subgroup analysis, we explored differences in cost-effectiveness between the type of maternity unit (SCU/HDU or NICU).

## RESULTS

Of the 155 maternity units in England, a total of 150 participated in the NPP. The five units that did not participate were pilot sites for the PReCePT QI toolkit development^11^ and were not included in the analysis (mean uptake of 98% in first half of 2020). From the 150 NPP units, a further 13 units were excluded as these comprised the PReCePT trial ESP group.^13^ This left 137 maternity units in the evaluation.

The NPP adoption dates of the participating units ranged from October 2018 to October 2020, although almost all units had started by April 2019. On average, there were 2.9 preterm births per unit per month. Maternal and baby characteristics were similar between the pre-implementation 12 months and post-implementation 12 months (Table 1). The average MgSO_4_ uptake across all units in the 12-month pre-implementation period was 70.9, increasing to 83.1% across the 12-month post-implementation period (Table 2). The average amount of missing MgSO_4_ data dropped from 2.9% to 1.4% across the comparison periods.

**Table 1:** The socio-demographic and clinical characteristics of mothers and babies in pre-term births in NPP maternity units in England, October 2017 – June 2020.

| Variable | Pre-implementation<sup>0</sup> |  | Post-implementation<sup>0</sup> |  |
| --- | --- | --- | --- | --- |
| Socio-demographic characteristics of babies<sup>1*</sup> |  |  |  |  |
| Number of babies (N) | 3,630 |  | 3,441 |  |
| Gestational age (weeks, median, IQR) | 27.9 | 25.9-29.0 | 27.9 | 26.0-29.1 |
| Birthweight (grams, median, IQR) | 982 | 770-1210 | 980 | 769-1205 |
| Birthweight adjusted for gestational age (z-score, median, IQR) | 0.1 | -0.6 – 0.7 | 0.1 | -0.6 – 0.7 |
| Male sex (N, %) | 1,960 | 54.0% | 1851 | 53.8% |
| Multiple births (N, %) | 871 | 24.0% | 817 | 23.7% |
| Socio-demographic and clinical characteristics of mothers<sup>2*</sup> |  |  |  |  |
| Number of mothers (N) | 3,189 |  | 3,016 |  |
| Maternal age (years, mean, SD)) | 31 | 6 | 31 | 6 |
| White ethnicity (N, %) | 1711 | 60.2% | 1610 | 61.2% |
| Level of deprivation (IMD quintile, N, %) |  |  |  |  |
| 1 (most deprived) | 1108 | 31.0% | 1112 | 32.9% |
| 2 | 912 | 25.6% | 773 | 22.9% |
| 3 | 643 | 18.0% | 621 | 18.4% |
| 4 | 485 | 14.0% | 454 | 13.4% |
| 5 (least deprived) | 422 | 11.82% | 418 | 12.4% |
| Hypertension in pregnancy (N, %) | 128 | 3.5% | 157 | 4.6% |
| Antenatal steroids given (N, %) | 3340 | 92.1% | 3220 | 93.9% |
| Maternity Unit characteristics |  |  |  |  |
| Special Care Unit / High Dependency Unit (N, %) | 1336 | 36.% | 1226 | 35.6% |
| Neonatal Intensive Care Unit (N, %) | 2294 | 63.2% | 2215 | 64.4% |
| Average number of eligible births per hospital per month (Mean, SD) | 2.9 | 2.1 | 2.9 | 2.1 |

<sup>0</sup> Figures cover the 12 months prior to, and 12 months following the recorded NPP adoption date at each unit, excluding the month of adoption itself.

**Table 2:** MgSO_4_ uptake in NPP maternity units in England October 2017 – June 2020^0^.

| Variable | Pre-implementation<sup>1</sup> |  | Post-implementation<sup>1</sup> |  |
| --- | --- | --- | --- | --- |
|  | N | % | N | % |
| Total number of eligible births | 3172 |  | 3014 |  |
| Total number of mothers **given** MgSO<sub>4</sub> | 2279 | 71.9% | 2527 | 83.8% |
| Total number of mothers **not given** MgSO<sub>4</sub> | 803 | 25.3% | 447 | 14.8% |
| Total number with MgSO<sub>4</sub> data **missing** | 90 | 2.8% | 40 | 1.3% |
| Mean MgSO<sub>4</sub> uptake across all units <sup>2</sup> | 70.9% | SD: 3.6% | 83.1% | SD: 3.5% |
| Mean MgSO<sub>4</sub> missing data across all units | 2.9% | SD: 1.3%<br>Range: 1.1% to 5.6% | 1.42% | SD: 1.0%<br>Range: 0% to 3.1% |
| Reason MgSO<sub>4</sub> not given |  |  |  |  |
| Contraindicated | 9 | 1.2% | 6 | 1.3% |
| Declined | 7 | 0.7% | 3 | 0.7% |
| Delivery imminent | 499 | 62.1% | 337 | 75.4% |
| Not appropriate | 69 | 8.6% | 24 | 5.4% |
| Not offered | 129 | 16.1% | 51 | 11.4% |
| Data missing | 90 | 11.2% | 26 | 5.8% |

<sup>0</sup> MgSO<sub>4</sub> data from records on singleton births and the first-born of multiples with records in the dataset.
<sup>1</sup> Covers the 12 months prior to, and following the recorded NPP adoption month at each unit
<sup>2</sup> Uptake percentages exclude missing data from the denominator.

Imminent delivery was the most common reason for why MgSO_4_ was not given (62.1% pre-implementation and 75.4% post-implementation). Pre-implementation, MgSO_4_ was recorded as ‘not offered’ in 16.1% of cases and post-implementation this had dropped to 11.4% (Table 2).

Overall, the trend in mean MgSO_4_ uptake increased steadily (Figure 1, supplementatry file - MgSO_4_ uptake through time) from October 2017 (earliest pre-implementation date) to June 2020 (the latest follow-up date). Average MgSO_4_ uptake varied by AHSN, and within each AHSN, high monthly variation in uptake was observed (supplementatry file - MgSO4 uptake by AHSN). The lowest average uptake was around 65% at the end of 2017, and the highest was around 94% around May 2020.

**Figure 1.**
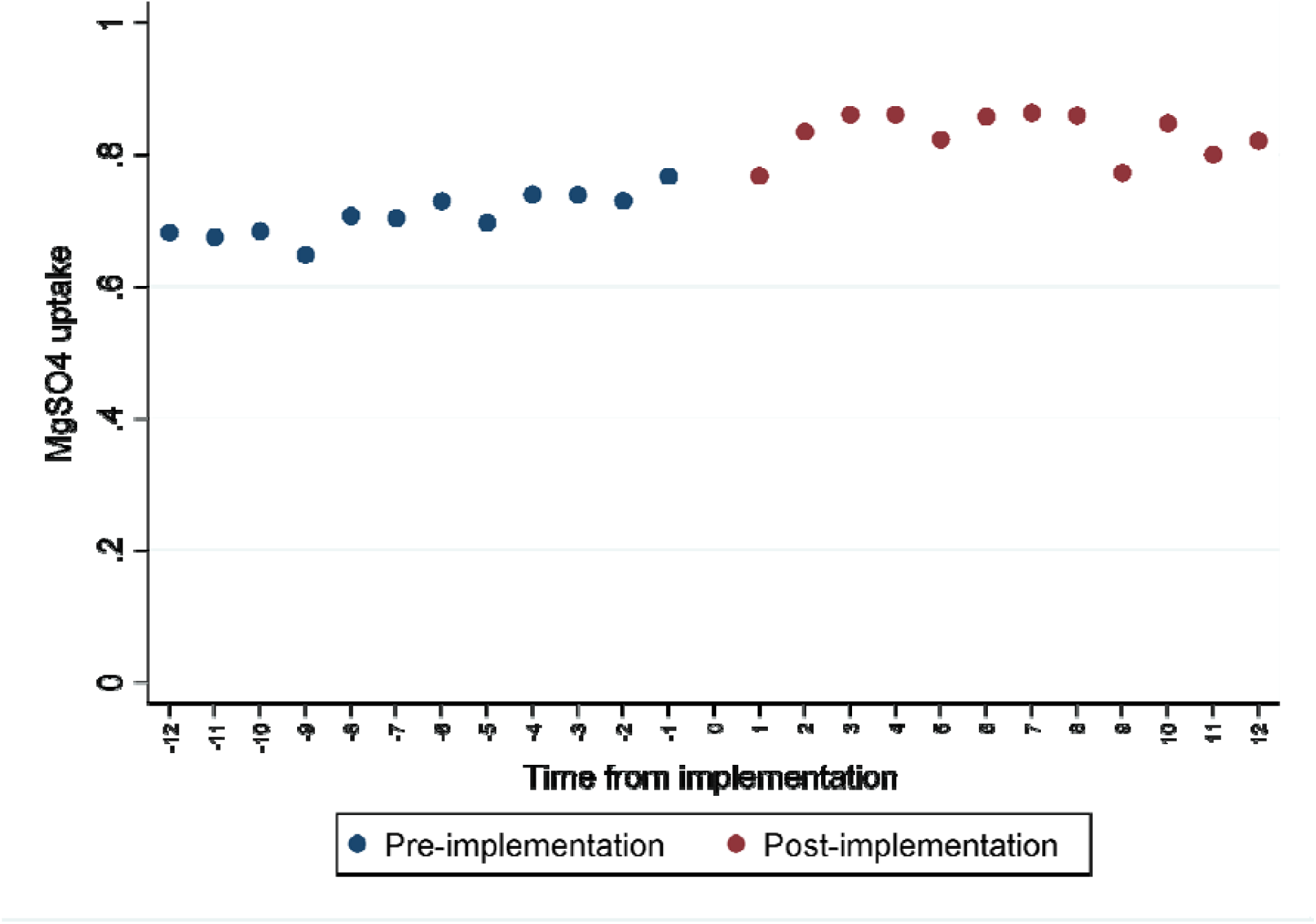
MgSO_4_ uptake pre- and post-implementation.

The unadjusted average increase in uptake between the pre- and post-implementation periods was 12.2 percentage points. After adjusting for unit size, clustering by AHSN, maternal, baby, and maternity unit factors, and time, the average increase in uptake between the pre- and post-implementation periods was 6.3 percentage points (95% CI 2.6 to 10.0 percentage points, p<0.001) (Table 3). After taking into account the likely variation in activities around the NPP adoption month (by removing from the analysis data for 3 months before and after the adoption date), the estimate changed to 9.5% (95% CI 4.3 to 14.7%, p<0.001) (Table 3).

**Table 3:** The difference in MgSO_4_ uptake after implementation of the NPP in maternity units in England.

|  | **Models** | **Difference in MgSO<sub>4</sub> uptake (percentage points)<sup>0</sup>** | **95% CI (percentage points)** | **p-value** |
| --- | --- | --- | --- | --- |
| 1 | Unadjusted<sup>1</sup> | 12.2 | 9.5 to 15.0 | <0.001 |
| 2 | Adjusted for unit size<sup>2</sup> | 11.0 | 8.9 to 13.1 | <0.001 |
| 3 | Adjusted for unit size and clustering by AHSN<sup>3</sup> | 11.0 | 8.4 to 13.5 | <0.001 |
| 4 | Adjusted for unit size, NPP month and clustering by AHSN<sup>4</sup> | 6.7 | 2.8 to 10.5 | 0.001 |
| 5 | Fully adjusted<sup>5</sup> | 6.3 | 2.6 to 10.0 | 0.001 |
| 6 | Fully adjusted model<sup>5</sup> + excluding records within 3 months either side of the start month | 9.5 | 4.3 to 14.7 | <0.001 |
| **Additional analyses** |  |  |  |  |
| 7 | Model 6 + including the 13 PReCePT trial intervention units | 9.6 | 4.4 to 14.8 | <0.001 |
| 8 | Model 6 + excluding units in one AHSN where implementation was delayed | 10.0 | 3.9 to 16.0 | 0.001 |

<sup>0</sup> Percentage point changes, post- minus pre-implementation.
<sup>1</sup> Crude regression of uptake post-implementation compared with pre-implementation.
<sup>2</sup> As per model 1, plus additionally weighted on number of eligible records per unit, with robust standard errors.
<sup>3</sup> As per model 2, plus additionally accounting for clustering by AHSN, with robust standard errors.
<sup>4</sup> As per model 3, plus additionally adjusted for recorded start month
<sup>5</sup> As per model 4, plus additionally adjusted for birthweight adjusted for gestational age and sex, maternal age, IMD, ethnicity, multiple birth, maternal hypertension (all unit-level aggregates), level of unit, study month

Neither including the PReCePT RCT study units nor excluding units from the one AHSN that started later than the others from the analysis had any substantial effect on the difference in uptake between pre-to post-implementation periods.

The proportion of missing MgSO_4_ data fluctuated during the NPP period between 0-7%, but overall tended to decrease over time (supplementary file - Trend in missing MgSO_4_ data over time). Around April 2020, at the start of the Covid pandemic, there appeared to be an increase in the amount of missing MgSO_4_ data.

### Costs and cost-effectiveness analyses

The mean funded implementation cost of the NPP was £6,044 per unit, £738 relating to national programme management, £2,764 funded support from AHSNs, and £2,500 funded backfill of clinical time for midwives. The mean implementation cost per baby was £267 per preterm baby born. The incremental impact of the NPP on uptake of MgSO_4_ during the 12 months follow-up was an absolute increase of 6.0 percentage points (95% CI 4.6% to 10.9%, p=0.03).

From a societal perspective, the NPP was associated with a mean increase of 0.01 QALYs per preterm baby and £649 total incremental savings over a baby’s lifetime. This equates to a net monetary benefit of £886 per preterm baby exposed to MgSO4 at a willingness-to-pay threshold of £20,000, and the probability of the NPP being cost-effective was greater than 95% (Table 4; supplementary file - Cost-effectiveness plane of National PreCePT Programme).

**Table 4:** Probabilistic Analysis Results of the NPP cost-effectiveness.

| **NPP** | **Combined** |  |  | **SCU/HDU units** |  |  | **NICU units** |  |  |
| --- | --- | --- | --- | --- | --- | --- | --- | --- | --- |
|  | Point estimate | Lower 95% limit | Upper 95% limit | Point estimate | Lower 95% limit | Upper 95% limit | Point estimate | Lower 95% limit | Upper 95% limit |
| **Incremental implementation costs, £** | 294 | 32 | 808 | 553 | 134 | 1,277 | 98 | 57 | 149 |
| **Incremental lifetime costs, £** | -943 | -1,486 | -469 | -909 | -1,514 | -362 | -867 | -1,443 | -357 |
| **Incremental total costs, £** | -649 | -1,284 | 35 | -356 | -1,131 | 539 | -769 | -1,344 | 258 |
| **Incremental QALYs** | 0.01 | 0.01 | 0.02 | 0.01 | 0.01 | 0.02 | 0.01 | 0.01 | 0.02 |
| **Net Monetary Benefit\*, £** | 886 | 130 | 1,621 | 586 | -372 | 1,475 | 987 | 355 | 1,675 |

\*We used a willingness-to-pay threshold of £20,000 per QALY

Although the NPP implementation cost per preterm baby was higher in small (SCU/HDU) units than the larger NICU units (Table 4), the probability of the NPP being cost-effective in small units was still high, at about 85% (supplementary file - Cost-effectiveness plane of National PreCePT Programme).

## DISCUSSION

### Summary of results

To our knowledge, this is the first evaluation of the impact of the implementation of a national perinatal QI programme focused on increasing administration of an evidence-based drug intervention in the UK. The principal finding was that the NPP increased the proportion of women who received MgSO_4_ for preterm births.

The NPP was cost-effective. From a societal lifetime perspective, the additional increase in MgSO_4_ use as a result of the NPP-generated improvements in health and cost savings. This beneficial impact of the NPP was also evident in smaller SCU/HDU units with comparatively high implementation costs per preterm baby. The NPP generated a net monetary benefit to society of approximately £3m over the 12 months following implementation.

The reduction in amount of missing MgSO_4_ data indicates an improvement in MgSO_4_ record-keeping. This is in line with the historical trend of gradually decreasing levels of missing MgSO_4_ data.

### Comparison with the literature

The uptake of MgSO_4_ varied across countries, with estimates of 0%-12.3% in Europe (2011-2015),^21^ and 43.0% in Canada (2011-2015).^22^ We found only one similar quality improvement strategy, MAG-CP, implemented in tertiary perinatal centres in Canada, which included educational rounds, focus group discussions and surveys of barriers and facilitators, on top of a national guideline and an online e-learning module.^22^ The MAG-CP intervention was associated with an absolute increase in uptake of 44.3%, from 2.0% in the pre-implementation period (2005-2010) to 46.3% after implementation (2011-2015).^22^

### Strengths and limitations

The study made use of routinely collected maternal and neonatal data from the NNRD and the NPP, providing robust, objective and high-quality data for the evaluation. This enabled the evaluation to cover all maternity units in England with minimal costs, making results generalisable We were able to obtain reliable NPP implementation cost data.

The use of mixed-effects models enabled comparisons of the MgSO_4_ uptake in the pre- and post-implementation periods while allowing adjustment for known potential confounders and clustering by AHSN. As implementation of the NPP is directed by the AHSNs, unmeasured similarities and differences between units within AHSNs need to be taken into account.

Our study has limitations. In a pragmatic before-and-after observational evaluation of this kind it is impossible to conclusively attribute the observed increase in uptake to the NPP alone. However, the statistical methods used in this evaluation minimise the impact of known biases and confounders, which gives reason to believe that the NPP did have a positive impact on uptake. The RCT comparing MgSO4 uptake in NPP units versus those receiving enhanced support is reported separately, but results were comparable to those reported here.^13,14^ The analyses are also limited to the available data (i.e., to June 2020), it is probable that NPP benefits will persist; therefore, sustainability analyses need to be addressed in future studies.

The NNRD dataset only includes records for live babies admitted to a registered neonatal unit; any baby who did not survive before admission to a neonatal unit is not recorded. In addition, the adoption dates used to demarcate the exposure periods were not firmly defined. NPP activities were reported to have started before or after the date stipulated by the AHSNs, possibly diluting the effects of the NPP on the uptake. Despite this, the various relevant sensitivity analyses did not alter the main findings.

The estimated changes have shown the average increase between the two periods, taking into account the variability between units and across time. The percentage point change after adjustment for possible confounding variables was smaller than expected based on previously audited estimates.^23^ This suggests that other factors such as organisational context and other structural enablers, as suggested in literature,^24^ could have contributed to the observed increase in the MgSO_4_ uptake, aside from the NPP. From our four year post-implementation experience in the five pilot sites, the improved administrator rate of MgSO_4_ is likely to be sustained, therefore, the study may underestimate the cost-effectiveness of the NPP.

Data showed a slight decrease in uptake of MgSO_4_ around April 2020, and an increase in the amount of missing MgSO_4_ data at the same time. These may simply be random fluctuations but are also consistent with a possible impact of the first wave of COVID-19 in England. The two could be associated given that the staffing pressures of a pandemic are likely to affect quality of care. Also, women may have presented at hospital later during this time due to caution about contact, meaning some missed opportunities to give MgSO_4_ due to imminent delivery. Further analysis of future data would be valuable to identify whether there are any trends in uptake or missing data associated with the course of the pandemic.

### Implications for clinical practice

Our study reinforces that active implementation of national initiatives utilising QI toolkits, clinical leadership, and coordinated by regional knowledge mobilis, ation teams (AHSNs) can have a substantial effect on accelerating uptake of evidence based therapies. Slow uptake comes at a clinical and economic cost with health gains foregone due to delay in the implementation of new innovations. Uptake of new evidence or guidelines is generally slow due to practical barriers, lack of knowledge and need for behaviour change. It is important to anticipate potential need for active implementaion to accelerate uptake and minimise this potential loss.

In conclusion, the study provides evidence that this quality improvement intervention was effective and cost-effective in increasing MgSO_4_ uptake in maternity units across England, and that successful national scale-up and implementation can be achieved through regional support. The NPP generated a net monetary benefit to society of approximately £3m over the 12 months following implementation, with potential to grow if MgSO4 administration rates are maintained or increased further.

## Data Availability

Anonymised individual-level data for this study comes from the NNRD. Our data sharing agreement with the NNRD prohibits sharing data extracts outside of the University of Bristol research team. The NNRD data dictionary is available online and copies of the Statistical Analysis Plan are available on available on the University of Bristols institutional repository (https://research-information.bris.ac.uk/en/projects/national-precept-prevention-of-cerebral-palsy-in-pre-term-labour-).

## Acknowledgements

We would like to thank all the participants who contributed to this evaluation. We also acknowledge the support of many staff in the AHSN Network and Brent Opmeer for their support and guidance.

## Ethics and regulatory considerations

The UK National Health Service Health Research Authority (HRA project ID: 260504) and the University of Bristol Faculty of Health Sciences Research Ethics Committee (FREC Ref: 84582) approved the conduct of the study.

## Transparency statement

KL affirms that the manuscript is an honest, accurate, and transparent account of the study being reported, that no important aspects of the study have been omitted, and that any discrepancies from the study as originally planned (and, if relevant, registered) have been explained.

## Contributors

KL, JD, NS and EW conceptualized the evaluation; KL and JM are Chief Investigators; TP, KT and MTR are quantitative evaluation leads; SR is process evaluation lead; WH and HM are health economic evaluation leads; PC and EMH are study managers; EW is NPP manager; ET, EW, NS and GF advised on the study methodology, implementation and analysis; HE, MTR, RM, CSR, EW and PC acquired NNRD, NPP and cost data; HE, MTR and RM conducted the effectiveness analysis; CSR, HM and WH conducted the cost-effectiveness analysis; HE, MTR and CSR wrote the original manuscript and contributed equally to the paper; all authors reviewed and edited the manuscript for content and approved the submission.

## Conflicts of interest

GF received grant funding from the NIHR and the British Heart Foundation and is a party to partnership agreements with industry partners as CEO of the Oxford Academic Health Science Network. He is chair of the Buckinghamshire, Oxfordshire and West Berkshire Integrated Stroke Delivery Network, the Academic Health Science Network, European Stroke Organisation Council of Fellows and the Academy of Medical Sciences Fellowship Sectional Committee 7. He is Director of the Cogentis and Accelerate companies, Non-Executive Director of the National Institute for Health and Care Excellence, serves on the Board of Trustees of the Picker Institute and Health Services Research UK, and the governing body of Green Templeton College, Oxford University. He is data monitoring committee member for the PREVENT-SVD study, trial steering group member for OPTIMAS, R4VaD, ATTEST-2 and SENIOR-RITA trials, grants review panel member for Pfizer/Bristol Myers Squibb, and round table member for the Bristol Myers Squibb/Price Waterhouse Cooper Life Sciences 2030 Cancer Moonshot and Astellas Company Conference.

KT acted as Expert Witness to the High Court in England, called by the UK MHRA, defendants in a case on hormonal pregnancy tests and congenital anomalies 2021/22

All other authors in this manuscript have no conflict of interest to declare aside from funding from NIHR ARC West, AHSN, NHS England and The Health Foundation as detailed above. We declare that the study management group have no competing financial, professional, or personal interests that might have influenced the study design or conduct.

## Public and Patient Involvement

Public and Patient Involvement for this study built on the involvement work in the PReCePT pilot study.^11^ This used a co-design and co-production approach including a partnership with BLISS, a support organisation for mothers experiencing pre-term births, and two mothers who had experienced pre-term births. These mothers were involved in study design

## Data sharing statement

Anonymised individual-level data for this study comes from the NNRD. Our data sharing agreement with the NNRD prohibits sharing data extracts outside of the University of Bristol research team. The NNRD data dictionary is available online^25^ and copies of the Statistical Analysis Plan are available on available on the University of Bristol’s institutional repository (https://research-information.bris.ac.uk/en/projects/national-precept-prevention-of-cerebral-palsy-in-pre-term-labour-).

