## Supplementary materials for "National PReCePT Programme: a quasi-experimental before-and-after evaluation of the implementation of a national quality improvement programme to increase the uptake of magnesium sulphate in pre-term deliveries"

**Supplementary Material**

### **National PReCePT Programme provisions**

| **Item** | **Definition** |
| --- | --- |
| PReCePT QI toolkit | Clinical guidance;  Pre-term labour proforma template;  Staff training presentations;  Parent leaflet;  Posters for display on the unit to raise staff awareness;  A QI Learning Log;  Project Dashboard;  Pens, magnets, lanyards and other aide-mémoires to promote MgSO_4_ to unit staff (if purchased) |
| QI training | Local level (AHSN or unit level) QI training and guidance to adapt materials for local use, cascaded from AHSN |
| Regional support | Support from a AHSN level clinical lead (obstetrician and neonatologist) and AHSN lead |
| Local clinical champion | Local obstetrician and neonatologist identified by unit to guide and oversee local implementation |
| Funded time for local midwife champion | Funded time of up to 90 hours per unit (on average 2 hours per week) |
| National support | AHSN Network steering group;  National lead and manager;  Shared learning events between AHSN managers leading the NPP in their area |

### **Table S1. Missing data on possible confounders, aggregated at maternity unit level**

| **Variable** | **Number missing** | **Proportion missing^0^** |
| --- | --- | --- |
| Birthweight | 2 | 0.03% |
| Multiple birth | 1 | 0.01% |
| Maternal age | 37 | 0.52% |
| Mother’s ethnicity | 1559 | 22.61% |
| Level of deprivation (IMD decile) | 123 | 1.74% |

^0^out of 7071 aggregated maternity unit level data points

### **MgSO_4_ uptake through time in maternity units in England**

##### **Figure S1. MgSO_4_ uptake through time in maternity units in England, October 2017 – June 2020**

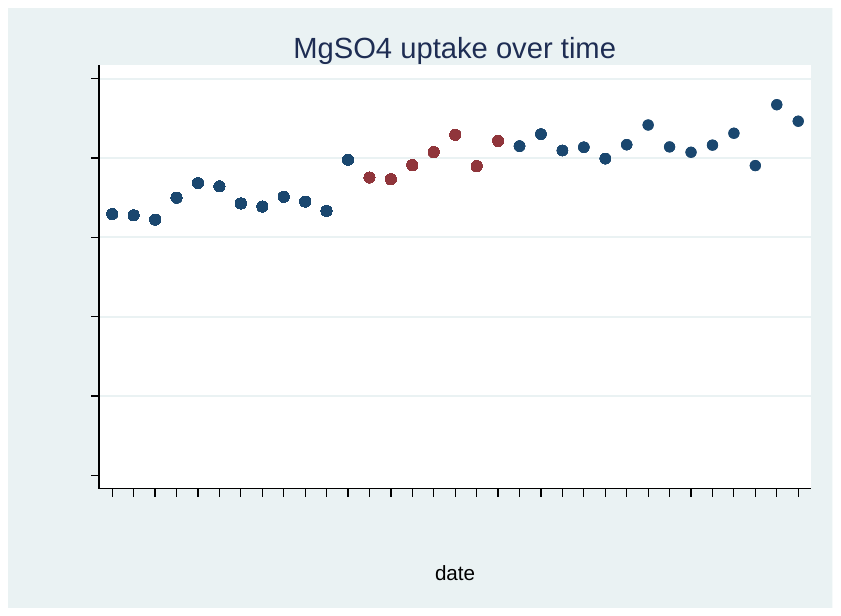

● Pre- or post-implementation periods ● NPP implementation start dates

##### **Figure S2. MgSO4 uptake through time in maternity units in England, January 2014 – June 2020**

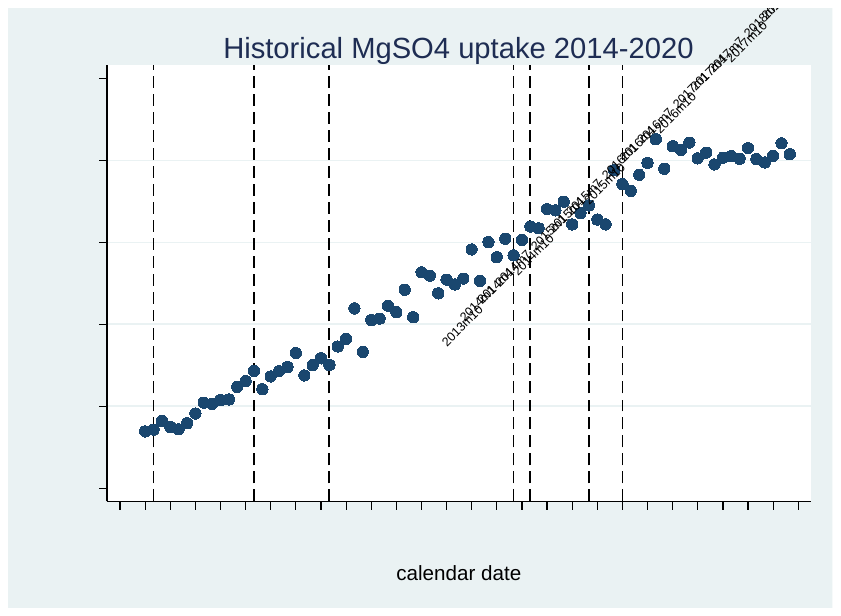

First NNAP Annual 🡪 Report

NICE Guidelines recommending MgSO4 in pre-term births

Publication

🡨 of PreCePT	Pilot Results

Start of NPP implementation

Wave 1🡪

Wave 2🡪

PReCePT Pilot Development and Implementation

##### **Figure S3. MgSO4 uptake by AHSN*, October 2017 – June 2020**

● Pre-implementation periods ● Post-implementation period (including NPP start date)

(a) **(b)**

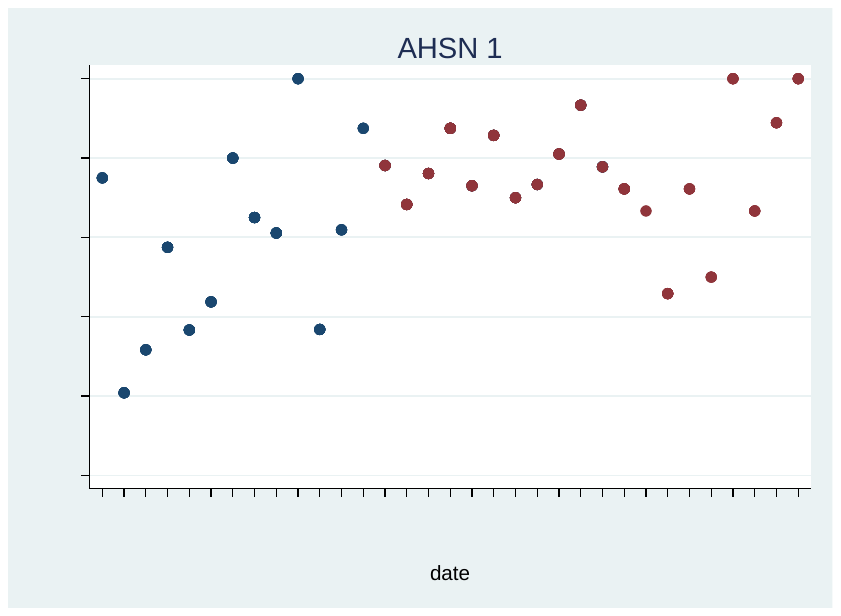

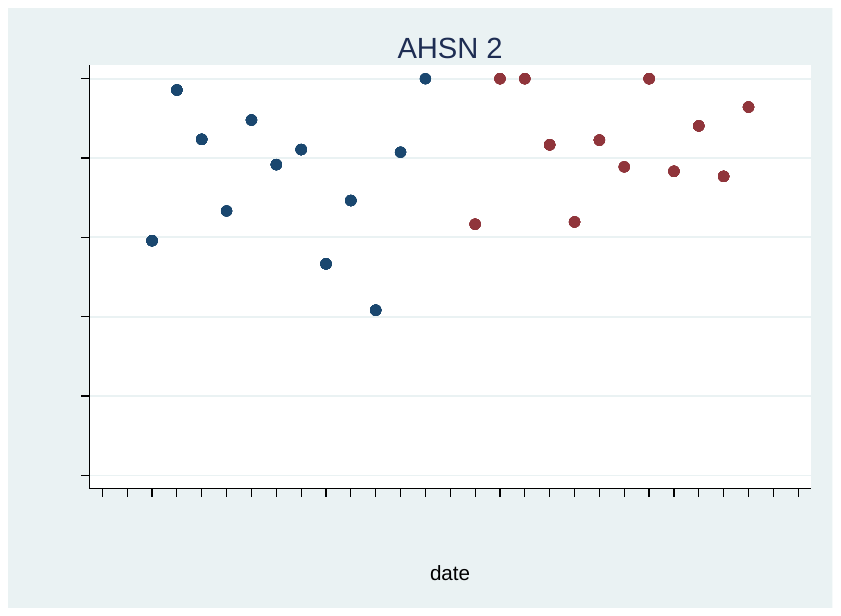

**(c)**	**(d)**

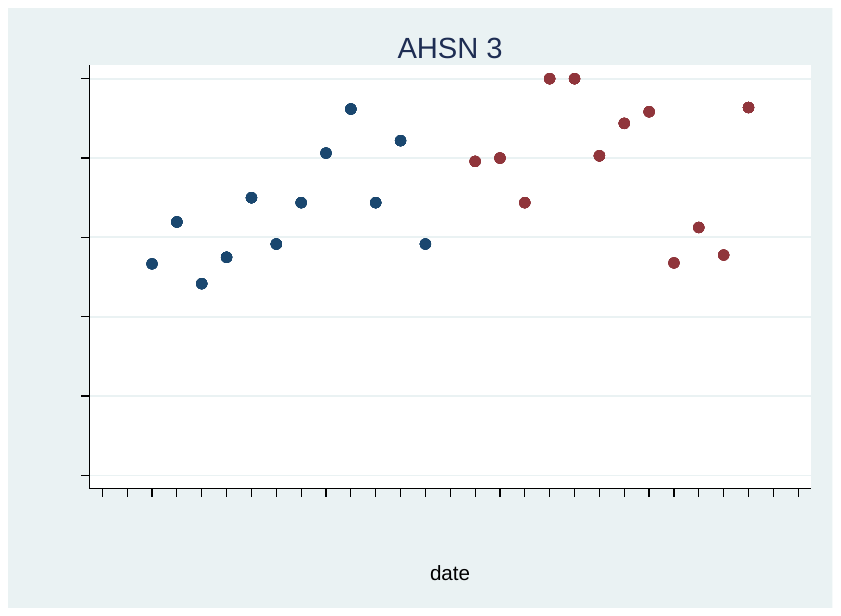

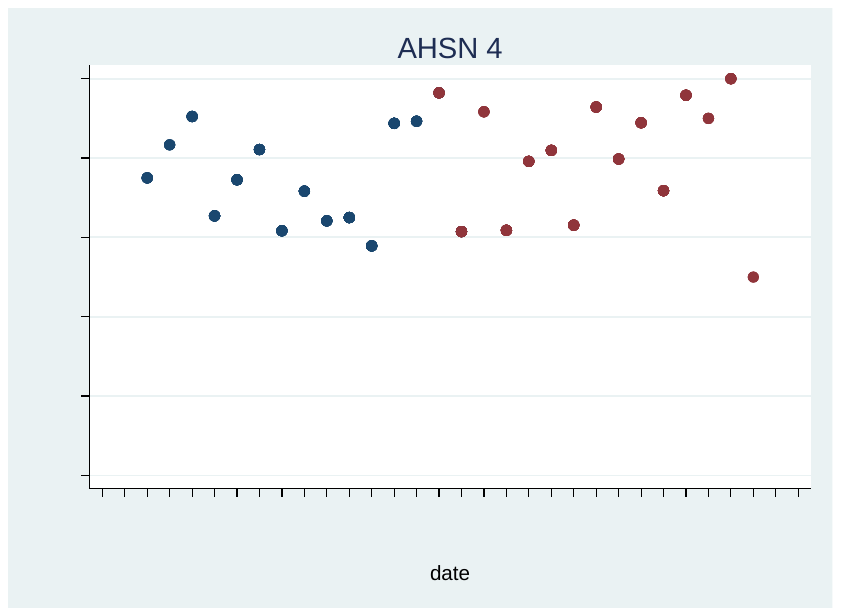

**(e)	(f)**

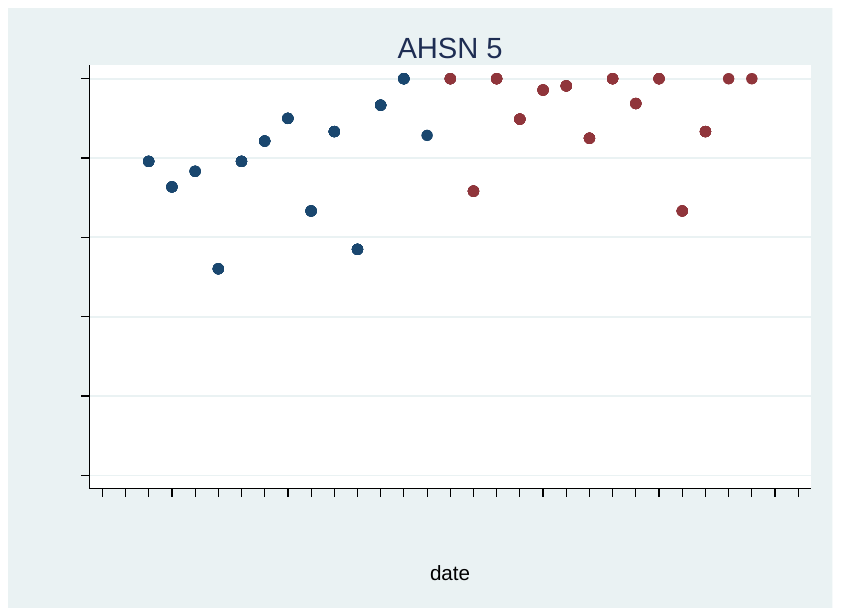

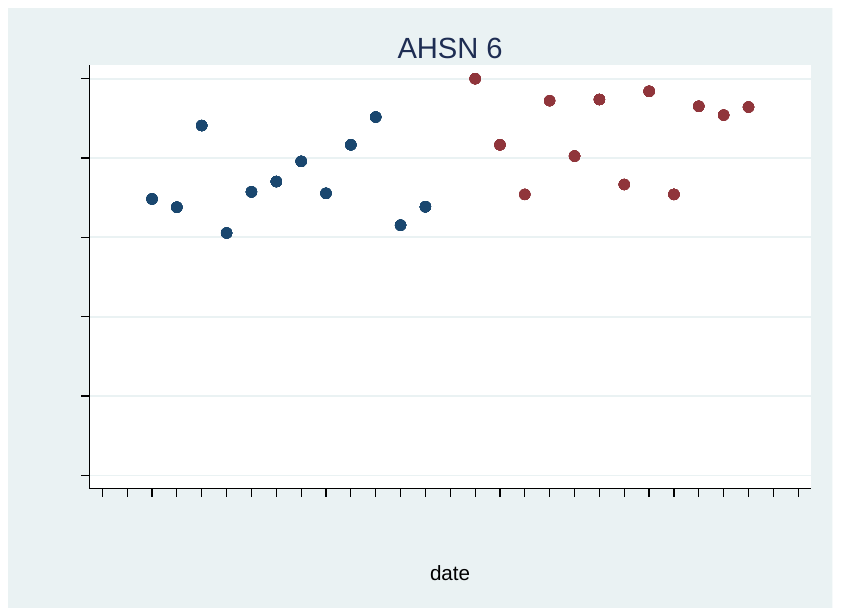

**Figure S3. MgSO_4_ uptake by AHSN*, October 2017 – June 2020 (cont.)**

● Pre-implementation periods ● Post-implementation period (including NPP start date)

**(h)	(i)**
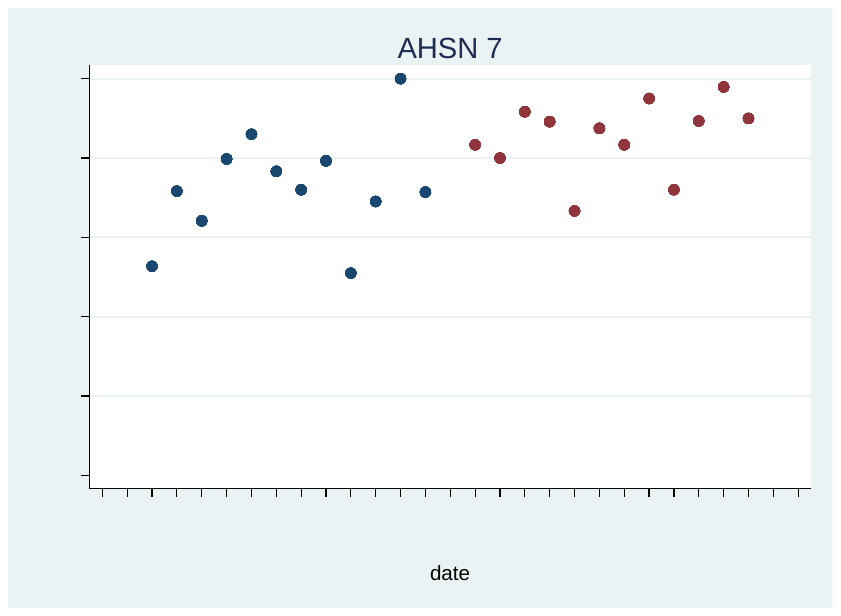

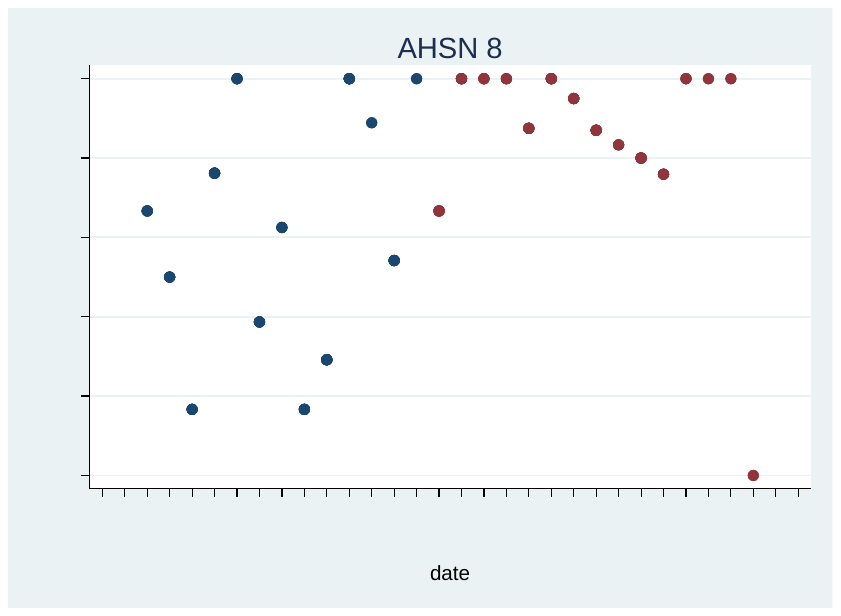

**(j)	(k)**

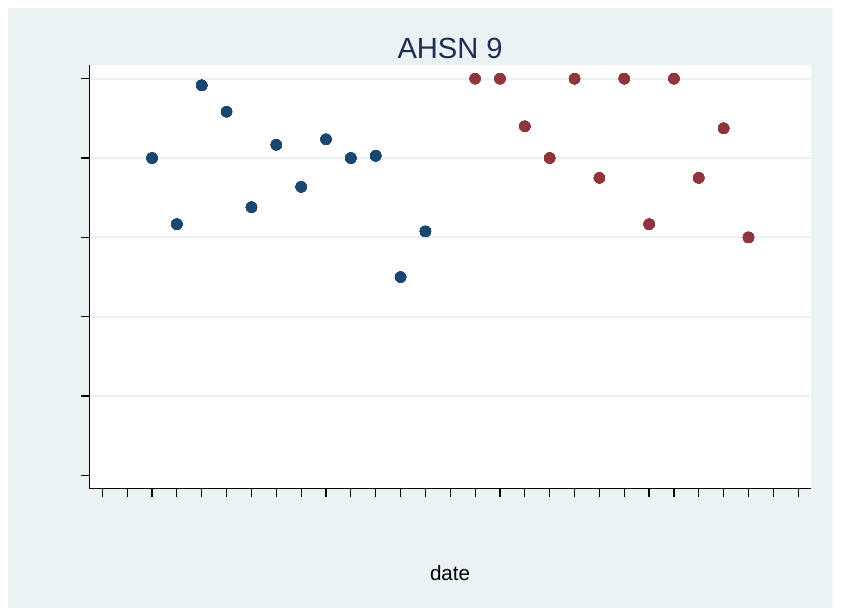

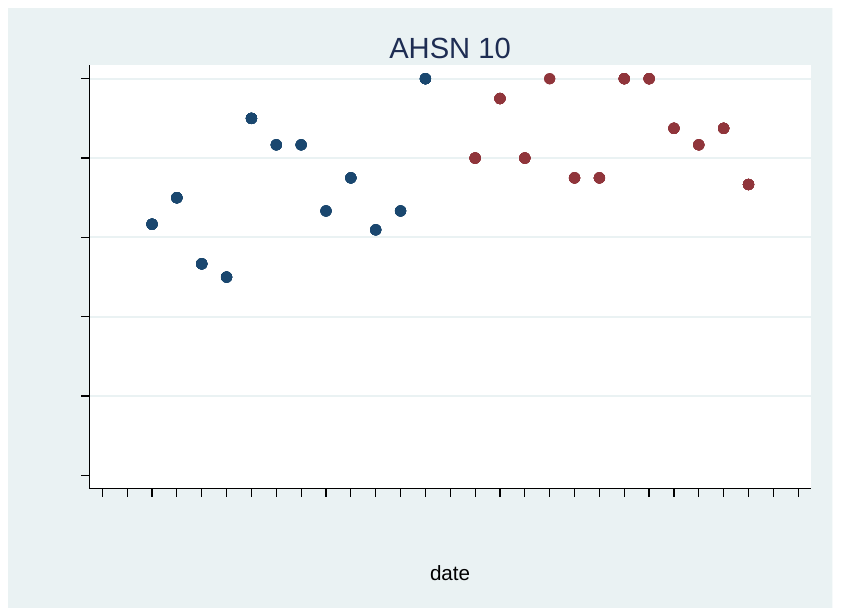

**(l)	(m)**

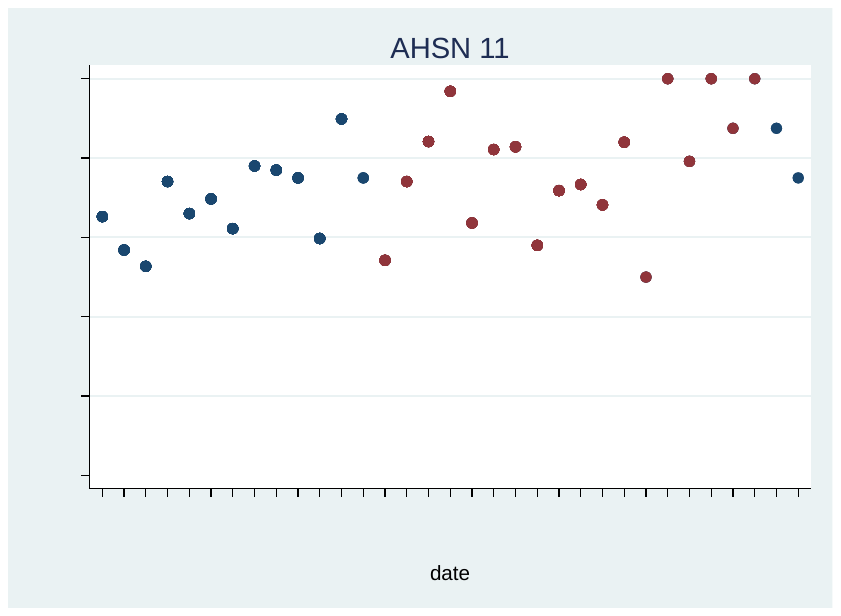

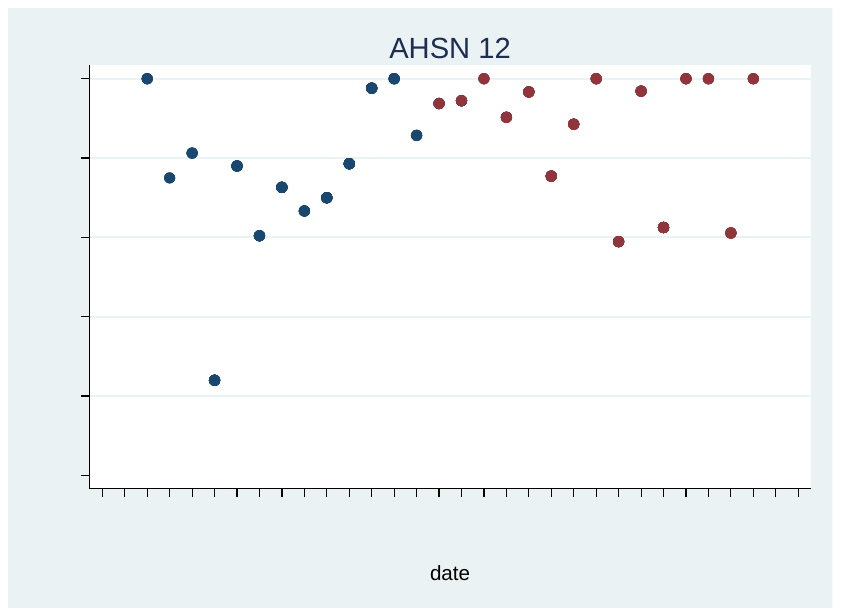

**Figure S3. MgSO_4_ uptake by AHSN*, October 2017 – June 2020 (cont.)**

● Pre-implementation periods ● Post-implementation period (including NPP start date)

**(n)	(o)**

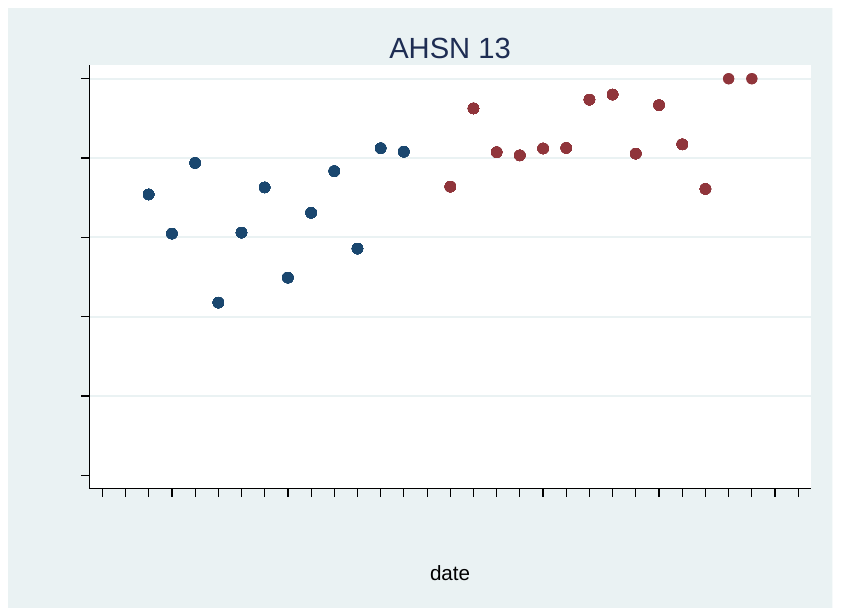

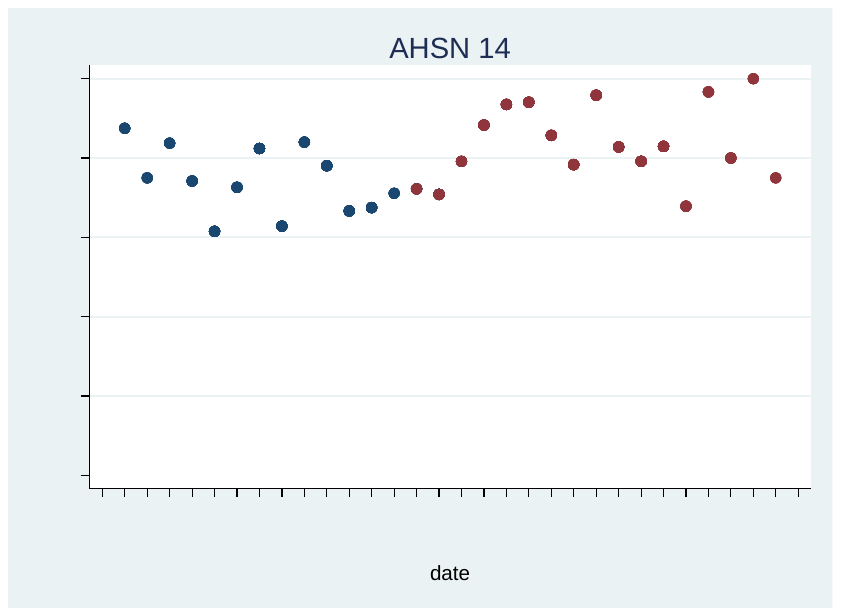

*One AHSN was comprised of pilot sites for the toolkit development and was not included in this study

### **Trend in missing MgSO_4_ data over time in maternity units in England**

##### **Figure S4. Trend in missing MgSO_4_ data over time, October 2017 – June 2020**

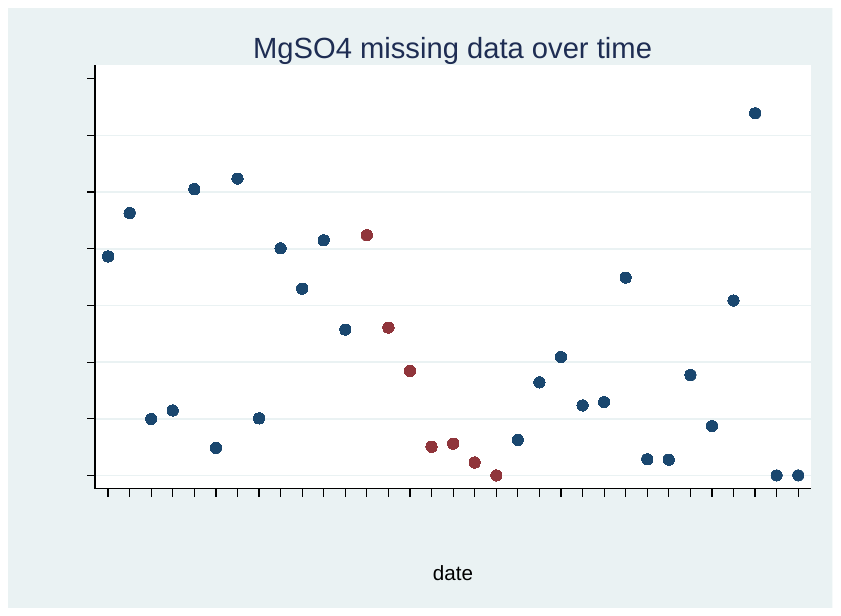

● Pre- or post-implementation periods ● NPP implementation start dates

##### **Figure S5. Trend in missing MgSO4 data over time, January 2014 – June 2020**

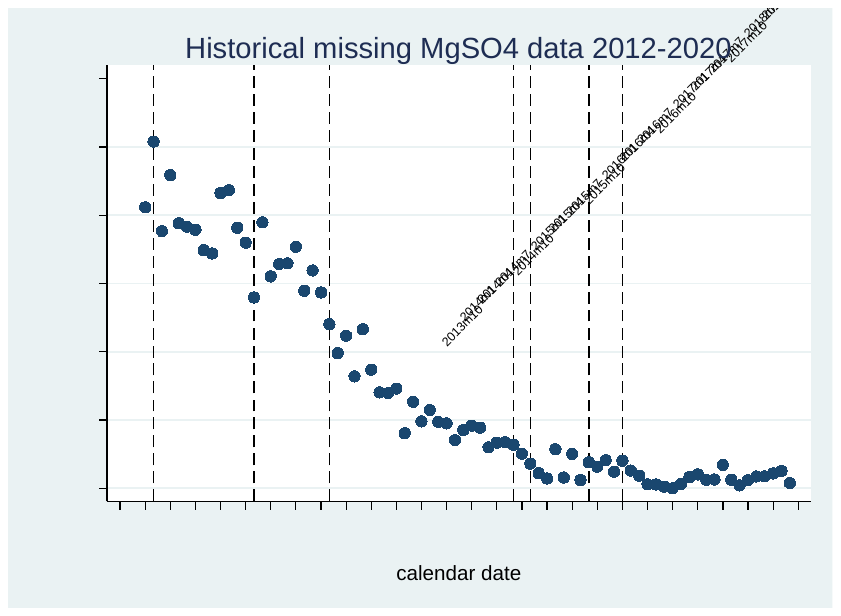

PReCePT Pilot Development and Implementation

NICE Guidelines recommending MgSO4 in pre-term births

🡨 Publication of PReCePT Pilot Results

First NNAP Annual 🡪 Report

Start of NPP implementation

Wave 1🡪

Wave 2🡪

### **Economic evaluation**

##### **Table S2. Estimated lifetime costs and QALYs per patient associated with MgSO4 treatment†**

| **Type of birth** | **Perspective** | **Method** | **Cost, £*** | **Δcost, £*** | **QALYs** | **ΔQALYs** |
| --- | --- | --- | --- | --- | --- | --- |
| Imminent | Societal | MgSO_4_ | 61,971 | -23,690 | 26.6 | 0.3 |
|  |  | No MgSO_4_ | 85,661 |  | 26.3 |  |
| Threatened | Societal | MgSO_4_ | 44,068 | -15,964 | 26.7 | 0.2 |
|  |  | No MgSO_4_ | 60,032 |  | 26.5 |  |

^†^ Based on Bickford et al (8)

^*^Cost estimates were converted to Pounds Sterling and inflated to 2019 prices

##### **Table S3. Point estimates, probability distributions, and source of parameter estimates used in the probabilistic analysis**

| **Parameter** | **Statistics** | **Combined Estimates** | **SCU/HDU Estimates** | **NICU Estimates** | **Probability distribution** | **Source** |
| --- | --- | --- | --- | --- | --- | --- |
| Type of birth |  |  |  |  |  |  |
| Imminent | n (%) | 971 (38%) | 336 (36%) | 635 (39%) | Beta distribution | NNRD data |
| Threatened | n (%) | 1573 (62%) | 598 (64%) | 975 (61%) | Beta distribution | NNRD data |
| Probability MgSO_4_ treatment (yes) – Baseline period | |  |  |  |  |  |
| MgSO_4_ yes - Imminent | n (%) | 679 (74%) | 228 (66%) | 451 (80%) | Beta distribution | NNRD data |
| MgSO_4_ yes - Threaten | n (%) | 1311 (73%) | 432 (64%) | 879 (78%) | Beta distribution | NNRD data |
| Effectiveness NPP |  |  |  |  |  |  |
| MgSO_4_ yes - Imminent | OR (se) | 1.09 (0.03) | 1.10 (0.04) | 1.07 (0.03) | LogNormal | Logistic regression |
| MgSO_4_ yes - Threaten | OR (se) | 1.05 (0.03) | 1.05 (0.04) | 1.04 (0.03) | LogNormal | Logistic regression |
| Health utility (QALY) |  |  |  |  |  |  |
| MgSO_4_ yes - Imminent | Mean (se*) | 0.3 (0.06) | 0.3 (0.06) | 0.3 (0.06) | Beta distribution | Bickford et al ^1^ |
| MgSO_4_ yes - Threaten | Mean (se*) | 0.2 (0.04) | 0.2 (0.04) | 0.2 (0.04) | Beta distribution | Bickford et al ^1^ |
| Lifetime costs |  |  |  |  |  |  |
| MgSO_4_ yes - Imminent | Mean (se*) | £-23,690 (-4,738) | £-23,690 (-4,738) | £-23,690 (-4,738) | Gamma distribution | Bickford et al ^1^ |
| MgSO_4_ yes - Threaten | Mean (se*) | £-15,964 (-3,193) | £-15,964 (-3,193) | £-15,964 (-3,193) | Gamma distribution | Bickford et al ^1^ |
| Implementation costs |  |  |  |  |  |  |
| MgSO_4_ yes - Imminent | Mean (sd) | £221 (341) | £473 (500) | £95 (44) | Gamma distribution | NNRD data |
| MgSO_4_ yes - Threaten | Mean (sd) | £325 (471) | £553 (701) | £99 (45) | Gamma distribution | NNRD data |
| MgSO_4_ no - Imminent | Mean (sd) | £259 (470) | £630 (596) | £97 (56) | Gamma distribution | NNRD data |
| MgSO_4_ no - Threaten | Mean (sd) | £360 (603) | £602 (765) | £106 (77) | Gamma distribution | NNRD data |

 *Standard Errors were assumed to be 20% if their point estimate

##### **Table S4. Mean lifetime QALYs and costs per baby by type of birth and trial arm**

|  | **Before NPP** | | | | | **Afer NPP** | | | | |
| --- | --- | --- | --- | --- | --- | --- | --- | --- | --- | --- |
|  | **Imminent** | | **Threatened** | | **Total** | **Imminent** | | **Threatened** | | **Total** |
|  | **MgSO_4_ Yes** | **MgSO_4_ No** | **MgSO_4_ Yes** | **MgSO_4_ No** |  | **MgSO_4_ Yes** | **MgSO_4_ No** | **MgSO_4_ Yes** | **MgSO_4_ No** |  |
| **Status distribution** |  |  |  |  |  |  |  |  |  |  |
| Combined | 28% | 10% | 45% | 17% | 100% | 31% | 7% | 47% | 15% | 100% |
| SCU/HDU units | 24% | 12% | 41% | 23% | 100% | 26% | 10% | 43% | 21% | 100% |
| NICU units | 31% | 8% | 47% | 13% | 100% | 34% | 6% | 49% | 11% | 100% |
| **Lifetime costs** |  |  |  |  |  |  |  |  |  |  |
| Combined | 61,971 | 85,661 | 44,068 | 60,032 | 55,917 | 61,971 | 85,661 | 44,068 | 60,032 | 54,982 |
| SCU/HDU units | 61,971 | 85,661 | 44,068 | 60,032 | 57,129 | 61,971 | 85,661 | 44,068 | 60,032 | 56,231 |
| NCIU units | 61,971 | 85,661 | 44,068 | 60,032 | 55,155 | 61,971 | 85,661 | 44,068 | 60,032 | 54,298 |
| **Implementation costs** |  |  |  |  |  |  |  |  |  |  |
| Combined | 0 | 0 | 0 | 0 | 0.00 | 221 | 325 | 259 | 360 | 267 |
| SCU/HDU units | 0 | 0 | 0 | 0 | 0.00 | 473 | 630 | 553 | 602 | 550 |
| NICU units | 0 | 0 | 0 | 0 | 0.00 | 95 | 97 | 99 | 106 | 98 |
| **QALYs** |  |  |  |  |  |  |  |  |  |  |
| Combine | 26.60 | 26.30 | 26.70 | 26.50 | 26.60 | 26.60 | 26.30 | 26.70 | 26.50 | 26.61 |
| SCU/HDU units | 26.60 | 26.30 | 26.70 | 26.50 | 26.58 | 26.60 | 26.30 | 26.70 | 26.50 | 26.59 |
| NICU units | 26.60 | 26.30 | 26.70 | 26.50 | 26.61 | 26.60 | 26.30 | 26.70 | 26.50 | 26.62 |

##### **Table S4. Deterministic Analysis Results of the NPP cost-effectiveness**

| **NPP** | **Combined** | **SCU/HDU units** | **NICU units** |
| --- | --- | --- | --- |
| Incremental implementation costs, £ | 267 | 550 | 98 |
| Incremental lifetime costs, £ | -934 | -897 | -857 |
| Incremental total costs, £ | -667 | -347 | -759 |
| Incremental QALYS | 0.01 | 0.01 | 0.01 |
| Net Monetary Benefit*, £ | 903 | 574 | 975 |

*We used a willingness-to-pay threshold of £20,000 per QALY

##### **Figure S6. Cost-effectiveness plane of National PreCePT Programme**

The graph displays results of Monte Carlo simulations with 10 000 iterations using the value ranges and distributions presented in Table S3. The horizontal axis represents the effect measures in Quality Adjusted Life Years (QALYs) for National PreCePT Programme; and the vertical axis represents the cost. Datapoints falling the top right quadrant indicate that the National PreCePT Programme is effective and costly. Datapoints falling bottom right quadrant indicate that National PreCePT Programme is effective and cost saving.

COMBINED

| QALYs, National PReCePT Programme | Cost, National PReCePT Programme |
| --- | --- |
| 0.00902493 | -658.195 |
| 0.0183221 | -1126.67 |
| 0.0096777 | 127.772 |
| 0.00755589 | -518.979 |
| 0.0142325 | -859.077 |
| 0.00691607 | 83.2929 |
| 0.00583017 | -283.365 |
| 0.0100978 | -774.423 |
| 0.00885982 | -731.599 |
| 0.0110851 | -1131.49 |
| 0.0101384 | -371.936 |
| 0.0138627 | -1218.37 |
| 0.00398482 | -231.12 |
| 0.0104037 | -825.836 |
| 0.014838 | -1195.69 |
| 0.00919595 | -368.685 |
| 0.0057761 | -314.507 |
| 0.00868531 | -643.765 |
| 0.013694 | -1204.67 |
| 0.00904118 | -678.559 |
| 0.00944627 | -784.995 |
| 0.0055268 | -572.523 |
| 0.0096472 | -510.243 |
| 0.00450075 | -227.895 |
| 0.0139227 | -611.142 |
| 0.0164881 | -1124.15 |
| 0.014822 | -422.222 |
| 0.0125412 | -1386.41 |
| 0.0124202 | -642.417 |
| 0.0173133 | -760.818 |
| 0.0125132 | -1196.42 |
| 0.011141 | -571.948 |
| 0.00593882 | -68.0886 |
| 0.00774559 | -509.739 |
| 0.00869866 | -749.571 |
| 0.0140452 | -390.622 |
| 0.009882 | -130.344 |
| 0.0153309 | -423.814 |
| 0.0143098 | -576.979 |
| 0.0108847 | -841.97 |
| 0.012359 | -696.125 |
| 0.0105839 | -1047.76 |
| 0.00976381 | -388.941 |
| 0.0141541 | -183.397 |
| 0.0115732 | -881.587 |
| 0.0124807 | -413.647 |
| 0.00508739 | -358.385 |
| 0.0144989 | -1060.38 |
| 0.0119339 | -1108.39 |
| 0.0137598 | -221.178 |
| 0.0165531 | -871.308 |
| 0.0108933 | -201.626 |
| 0.00970757 | -704.904 |
| 0.0136738 | -420.124 |
| 0.00642236 | -505.553 |
| 0.00549529 | 202.997 |
| 0.0192308 | -1175.87 |
| 0.0155285 | -1162.72 |
| 0.00557397 | 297.971 |
| 0.0122837 | -989.891 |
| 0.0119132 | -199.732 |
| 0.0142155 | -585.021 |
| 0.00508136 | 188.41 |
| 0.00970669 | -112.505 |
| 0.0168468 | -1001.84 |
| 0.0148169 | -532.726 |
| 0.014529 | -924.612 |
| 0.0135205 | -984.604 |
| 0.0110225 | -372.305 |
| 0.021554 | -1094.61 |
| 0.00816832 | -440.596 |
| 0.0152417 | -1380.15 |
| 0.0120104 | -1071.77 |
| 0.00940034 | 1050.88 |
| 0.00911008 | -846.265 |
| 0.0140964 | -1409.21 |
| 0.0087224 | -240.638 |
| 0.0251834 | -1372.38 |
| 0.0209213 | -1015.62 |
| 0.016121 | -1084.33 |
| 0.00951807 | -643.003 |
| 0.00916749 | -288.36 |
| 0.0131456 | -1236.19 |
| 0.00482722 | 39.0067 |
| 0.00775573 | -257.008 |
| 0.0105715 | -488.507 |
| 0.00637609 | 184.066 |
| 0.00941195 | -459.713 |
| 0.00768498 | -315.187 |
| 0.00719835 | -430.874 |
| 0.0124486 | -934.301 |
| 0.00948593 | -711.706 |
| 0.0126159 | -1056.78 |
| 0.00852733 | -224.505 |
| 0.00517665 | -522.947 |
| 0.0137781 | -988.741 |
| 0.0156704 | -1171.42 |
| 0.0170653 | -1547.03 |
| 0.0121211 | -762.027 |
| 0.010546 | 27.6357 |
| 0.0160428 | -902.928 |
| 0.0108598 | -454.811 |
| 0.00895812 | 235.637 |
| 0.00787542 | -389.887 |
| 0.0176573 | -1104.62 |
| 0.0063222 | 55.8918 |
| 0.0100728 | -512.545 |
| 0.00501202 | -243.031 |
| 0.00922415 | -315.481 |
| 0.015248 | -915.398 |
| 0.0107757 | -896.473 |
| 0.0258069 | -1306.01 |
| 0.012431 | -670.394 |
| 0.00896399 | -709.872 |
| 0.011931 | -591.01 |
| 0.0122825 | -660.635 |
| 0.0265844 | -2103.12 |
| 0.0113157 | -755.88 |
| 0.00906681 | -972.289 |
| 0.0114417 | -227.748 |
| 0.0101922 | -717.49 |
| 0.0150002 | -910.938 |
| 0.0124716 | -431.66 |
| 0.0120867 | -212.042 |
| 0.0124419 | -241.58 |
| 0.0153442 | -493.876 |
| 0.00756257 | -222.112 |
| 0.0103638 | -460.002 |
| 0.0147983 | -919.907 |
| 0.0188501 | -734.178 |
| 0.0141704 | -894.387 |
| 0.0114401 | -400.845 |
| 0.0157472 | -1548.77 |
| 0.0147939 | -863.345 |
| 0.0119066 | -829.093 |
| 0.0124654 | -655.389 |
| 0.021332 | -1415.47 |
| 0.0115844 | 84.7648 |
| 0.0182278 | -835.618 |
| 0.00807029 | -425.662 |
| 0.00900635 | -88.4349 |
| 0.00850544 | -22.3478 |
| 0.0129433 | -595.466 |
| 0.0123808 | -762.313 |
| 0.0057677 | -387.536 |
| 0.0191821 | -1173.05 |
| 0.0114552 | -514.058 |
| 0.0085834 | -610.635 |
| 0.00881827 | -263.603 |
| 0.016721 | -1095.47 |
| 0.0119474 | -1146.7 |
| 0.00478693 | -96.9736 |
| 0.0110951 | -18.7366 |
| 0.00954636 | -775.483 |
| 0.00577025 | -506.301 |
| 0.0127365 | -945.893 |
| 0.0116655 | -689.034 |
| 0.00747667 | 652.165 |
| 0.0062703 | -414.144 |
| 0.0152103 | -959.711 |
| 0.0113053 | -292.679 |
| 0.00758319 | -543.321 |
| 0.0164658 | -857.077 |
| 0.0133452 | -853.98 |
| 0.0163912 | -962.609 |
| 0.0123486 | -678.168 |
| 0.013857 | -1140.74 |
| 0.00935172 | -560.049 |
| 0.0119956 | -490.665 |
| 0.0103245 | 161.654 |
| 0.0142447 | -86.2473 |
| 0.0113674 | -356.399 |
| 0.0166051 | 60.2106 |
| 0.0128698 | -1177.78 |
| 0.0118291 | -854.118 |
| 0.0119999 | -593.893 |
| 0.0114952 | -42.9038 |
| 0.00582385 | 291.668 |
| 0.0109549 | -1057.8 |
| 0.0201219 | -889.553 |
| 0.015135 | -466.736 |
| 0.0078861 | -214.271 |
| 0.0209973 | -1097.56 |
| 0.0126977 | -814.527 |
| 0.0139902 | -538.127 |
| 0.00363671 | 133.753 |
| 0.00517367 | -235.324 |
| 0.0106497 | -487.322 |
| 0.0123793 | -577.228 |
| 0.0115835 | -710.511 |
| 0.0195387 | -851.181 |
| 0.00849702 | -999.432 |
| 0.00877247 | -657.536 |
| 0.00947594 | -643.955 |
| 0.0112549 | -913.756 |
| 0.0169089 | -1327.74 |
| 0.00980402 | -669.039 |
| 0.0104613 | -540.611 |
| 0.00978176 | -639.996 |
| 0.0080944 | -403.475 |

##### **Figure S7. Cost-effectiveness plane of National PreCePT Programme for SCU/HDU units**

The graph displays results of Monte Carlo simulations with 10 000 iterations using the value ranges and distributions presented in Table S3 for SCU/HDU units. The horizontal axis represents the effect measures in Quality Adjusted Life Years (QALYs) for National PreCePT Programme; and the vertical axis represents the cost. Datapoints falling the top right quadrant indicate that the National PreCePT Programme is effective and costly. Datapoints falling bottom right quadrant indicate that National PreCePT Programme is effective and cost saving.

SCU/HDU

| QALYs, National PReCePT Programme | Cost, National PReCePT Programme |
| --- | --- |
| 0.00876185 | 276.196 |
| 0.00880194 | -312.123 |
| 0.0263321 | 65.9259 |
| 0.0174265 | -885.771 |
| 0.0125039 | 28.5155 |
| 0.00161842 | 212.659 |
| 0.0200501 | -906.99 |
| 0.013833 | -634.724 |
| 0.0124633 | -346.293 |
| 0.00582834 | 128.397 |
| 0.0149558 | -484.783 |
| 0.0137542 | -1171.15 |
| 0.0211512 | -836.254 |
| 0.010605 | -414.884 |
| 0.0115219 | -758.672 |
| 0.00758159 | 169.28 |
| 0.00762031 | 188.413 |
| 0.0120788 | -739.318 |
| 0.0112547 | -549.946 |
| 0.0154467 | -569.704 |
| 0.0123915 | -430.329 |
| 0.0107075 | -219.402 |
| 0.00285684 | 34.7619 |
| 0.0155556 | -739.887 |
| 0.0135725 | -522.54 |
| 0.0150856 | 335.562 |
| 0.00599509 | 82.7551 |
| 0.013256 | -920.637 |
| 0.0143042 | -601.214 |
| 0.0163538 | -493.567 |
| 0.00727105 | -47.3531 |
| 0.0142415 | -633.499 |
| 0.0161433 | -1157.75 |
| 0.00780441 | 567.602 |
| 0.00818391 | -829.333 |
| 0.0134142 | 563.538 |
| 0.0165674 | -1010.16 |
| 0.0194686 | 167.908 |
| 0.00950601 | -21.1762 |
| 0.0139785 | 178.613 |
| 0.0101974 | -619.849 |
| 0.0149791 | -194.409 |
| 0.00559581 | -76.5315 |
| 0.0132054 | -957.832 |
| 0.0142928 | -747.861 |
| 0.0118555 | 617.187 |
| 0.00525083 | -101.408 |
| 0.0154034 | -999.039 |
| 0.0111248 | 99.6861 |
| 0.0149121 | -723.88 |
| 0.0110013 | -232.846 |
| 0.0170461 | -189.871 |
| 0.00840669 | 897.288 |
| 0.0115668 | -689.235 |
| 0.0108617 | -533.099 |
| 0.0147635 | -1358.18 |
| 0.0127001 | -164.76 |
| 0.00847989 | -467.365 |
| 0.0224834 | -839.605 |
| 0.0145667 | -1017.79 |
| 0.0116628 | -396.88 |
| 0.00499457 | 361.39 |
| 0.0267327 | -1798.66 |
| 0.00769919 | -476.002 |
| 0.00836857 | -318.257 |
| 0.00669827 | -123.834 |
| 0.00822982 | -694.525 |
| 0.00923633 | -439.33 |
| 0.0139713 | -547.042 |
| 0.010142 | -496.189 |
| 0.00858085 | -215.97 |
| 0.0118481 | -606.022 |
| 0.0161513 | -1078.5 |
| 0.00836005 | -405.069 |
| 0.0220232 | -1215.5 |
| 0.00803177 | -379.219 |
| 0.00655549 | -176.762 |
| 0.00748252 | 553.714 |
| 0.0210156 | -825.818 |
| 0.0162063 | -926.599 |
| 0.00801209 | -178.503 |
| 0.0165757 | -958.027 |
| 0.0118613 | -498.507 |
| 0.0162502 | -533.448 |
| 0.0114275 | -608.522 |
| 0.00774044 | -535.677 |
| 0.0109669 | -391.186 |
| 0.00931314 | -202.506 |
| 0.0085081 | 16.88 |
| 0.00957813 | -678.79 |
| 0.0134918 | -660.04 |
| 0.0209841 | -1147.05 |
| 0.00693547 | 745.843 |
| 0.00807064 | -688.78 |
| 0.00862054 | -316.804 |
| 0.0174856 | -1094.52 |
| 0.0174972 | -1512.34 |
| 0.0124276 | -590.525 |
| 0.0217026 | -1202.21 |
| 0.00877475 | -457.122 |
| 0.014248 | -713.714 |
| 0.00822492 | 330.743 |
| 0.00652565 | -174.323 |
| 0.0173666 | -755.663 |
| 0.0121315 | 225.542 |
| 0.0147567 | -786.586 |
| 0.00434737 | 241.959 |
| 0.0185354 | -974.207 |
| 0.0108707 | -720.144 |
| 0.0124654 | -999.393 |
| 0.0109292 | -465.9 |
| 0.0249992 | -650.211 |
| 0.0135781 | -588.892 |
| 0.0122401 | -520.569 |
| 0.00970299 | -172.028 |
| 0.0114725 | 663.219 |
| 0.00836833 | -347.508 |
| 0.0172096 | -713.577 |
| 0.0131652 | -654.317 |
| 0.0162883 | 309.478 |
| 0.0114864 | -633.78 |
| 0.0202454 | -948.439 |
| 0.0119565 | -382.848 |
| 0.0154991 | -1063.34 |
| 0.00371224 | -70.9973 |
| 0.0179997 | -555.069 |
| 0.00915555 | 796.267 |
| 0.0116529 | -179.746 |
| 0.00829354 | -13.3144 |
| 0.00921023 | -324.178 |
| 0.00924706 | -672.341 |
| 0.0181794 | -917.734 |
| 0.00407243 | 1414.53 |
| 0.011459 | -535.129 |
| 0.0117181 | -956.196 |
| 0.00845201 | -100.533 |
| 0.0203456 | -1546.69 |
| 0.0136852 | -740.75 |
| 0.0178748 | -1032.71 |
| 0.0150783 | -1179.74 |
| 0.0207967 | -1184.73 |
| 0.0147313 | -1122.55 |
| 0.0144885 | -909.332 |
| 0.0104719 | -525.055 |
| 0.0095552 | 532.882 |
| 0.0140946 | -390.705 |
| 0.0113011 | -314.839 |
| 0.0091338 | -406.315 |
| 0.0155869 | -196.229 |
| 0.0153297 | -940.104 |
| 0.015078 | 469.495 |
| 0.0124202 | -271.415 |
| 0.0131425 | -437.118 |
| 0.0102095 | 200.447 |
| 0.00697132 | 365.056 |
| 0.0114776 | -407.16 |
| 0.00403025 | 1176.98 |
| 0.0100961 | -713.176 |
| 0.0137079 | -658.266 |
| 0.0190555 | -1273.64 |
| 0.016462 | -782.681 |
| 0.0137991 | -438.203 |
| 0.015927 | -836.575 |
| 0.0150442 | -541.337 |
| 0.00680023 | 73.2845 |
| 0.0112263 | -828.592 |
| 0.0135756 | -406.828 |
| 0.0103116 | -134.839 |
| 0.0118353 | 155.201 |
| 0.00929476 | -188.209 |
| 0.00867015 | 332.834 |
| 0.00908111 | -145.193 |
| 0.0114089 | -361.584 |
| 0.0107767 | 67.6831 |
| 0.0147046 | -658.274 |
| 0.008711 | -56.1446 |
| 0.0150501 | -861.252 |
| 0.0113407 | -349.258 |
| 0.00994037 | -752.937 |
| 0.020276 | -559.561 |
| 0.0112973 | -671.571 |
| 0.0122267 | -538.077 |
| 0.00894268 | 662.659 |
| 0.0184974 | -954.815 |
| 0.0170381 | -652.121 |
| 0.0149852 | -718.825 |
| 0.0133606 | -782.817 |
| 0.0118391 | -388.911 |
| 0.0144371 | -879 |
| 0.0143873 | -354.28 |
| 0.00885989 | -129.504 |
| 0.0100065 | -546.006 |
| 0.0156658 | -1040.67 |
| 0.00858377 | -270.112 |
| 0.0132798 | -346.863 |
| 0.00663937 | 562.387 |
| 0.0084208 | 56.5423 |
| 0.00706728 | 177.023 |
| 0.014241 | -737.859 |
| 0.00899298 | -579.019 |

##### **Figure S8. Cost-effectiveness plane of National PreCePT Programme for NICU units**

The graph displays results of Monte Carlo simulations with 10 000 iterations using the value ranges and distributions presented in Table S3 for NICU units. The horizontal axis represents the effect measures in Quality Adjusted Life Years (QALYs) for National PreCePT Programme; and the vertical axis represents the cost. Datapoints falling bottom right quadrant indicate that National PreCePT Programme is effective and cost saving.

NICU

| QALYs, National PReCePT Programme | Cost, National PReCePT Programme |
| --- | --- |
| 0.00866285 | -714.495 |
| 0.0166371 | -1001.45 |
| 0.0127361 | -1048.62 |
| 0.00634203 | -888.182 |
| 0.0118987 | -835.993 |
| 0.016886 | -1353.32 |
| 0.0158978 | -1487.28 |
| 0.00581096 | -457.148 |
| 0.00425948 | -106.673 |
| 0.00368073 | -134.723 |
| 0.00778813 | -669.289 |
| 0.0101816 | -606.931 |
| 0.0169281 | -646.342 |
| 0.0129315 | -1001.17 |
| 0.00999461 | -837.683 |
| 0.00877522 | -633.032 |
| 0.00894471 | -550.128 |
| 0.006575 | -589.844 |
| 0.0100174 | -734.418 |
| 0.00515413 | -264.866 |
| 0.00687923 | -707.904 |
| 0.0114093 | -622.426 |
| 0.010154 | -899.54 |
| 0.0112185 | -834.24 |
| 0.00627722 | -320.576 |
| 0.0111312 | -820.464 |
| 0.00859434 | -489.286 |
| 0.00408163 | -257.761 |
| 0.00695667 | -349.052 |
| 0.0164119 | -987.668 |
| 0.0146842 | -832.189 |
| 0.0109013 | -799.769 |
| 0.00875931 | -654.741 |
| 0.00669536 | -311.255 |
| 0.00446615 | -236.075 |
| 0.00695065 | -741.274 |
| 0.0100441 | -945.102 |
| 0.00818614 | -685.327 |
| 0.0113995 | -937.597 |
| 0.00413888 | -181.814 |
| 0.0106408 | -843.961 |
| 0.00968435 | -508.183 |
| 0.00933987 | -469.421 |
| 0.0120325 | -741.792 |
| 0.00614534 | -532.355 |
| 0.00265295 | 79.9425 |
| 0.00640379 | -370.039 |
| 0.0129005 | -1034.59 |
| 0.00817137 | -671.068 |
| 0.00379092 | -416.789 |
| 0.0138745 | -956.316 |
| 0.00955491 | -528.868 |
| 0.00916589 | -948.28 |
| 0.0117983 | -873.22 |
| 0.00961751 | -699.877 |
| 0.0161815 | -1148.31 |
| 0.0134116 | -892.192 |
| 0.0150148 | -1151.16 |
| 0.00765686 | -744.614 |
| 0.0133967 | -956.769 |
| 0.0161372 | -887.864 |
| 0.00986161 | -705.605 |
| 0.0140677 | -1118.51 |
| 0.0122833 | -771.515 |
| 0.0078719 | -606.275 |
| 0.00428567 | -484.258 |
| 0.0199124 | -992.59 |
| 0.00472021 | -242.527 |
| 0.00500428 | -359.922 |
| 0.00868319 | -565.995 |
| 0.0122982 | -869.192 |
| 0.000996582 | -4.88246 |
| 0.00759645 | -424.504 |
| 0.0102752 | -688.099 |
| 0.00889473 | -778.194 |
| 0.0114962 | -809.773 |
| 0.0150834 | -936.239 |
| 0.0154241 | -1395.04 |
| 0.0102702 | -591.146 |
| 0.0153366 | -865.995 |
| 0.00361093 | -144.028 |
| 0.0206739 | -1434.22 |
| 0.0155289 | -1106.98 |
| 0.00804569 | -586.743 |
| 0.00764004 | -421.231 |
| 0.0183342 | -1219.2 |
| 0.00751689 | -610.787 |
| 0.0163516 | -1279.2 |
| 0.0109095 | -821.581 |
| 0.00815624 | -488.886 |
| -0.0036211 | 379.799 |
| 0.0082417 | -335.628 |
| 0.00258863 | -155.282 |
| 0.00942482 | -705.102 |
| 0.00713265 | -635.84 |
| 0.0152344 | -982.97 |
| 0.0093036 | -672.718 |
| 0.0113184 | -942.557 |
| 0.010791 | -1253.59 |
| 0.0091768 | -720.658 |
| 0.0163344 | -1088.03 |
| 0.013486 | -752.651 |
| 0.0195789 | -1189.15 |
| 0.00962731 | -745.56 |
| 0.0164106 | -919.765 |
| 0.0102002 | -857.947 |
| 0.0130368 | -910.561 |
| 0.00834384 | -384.741 |
| 0.00957478 | -616.514 |
| 0.0108075 | -503.821 |
| 0.0170551 | -1166.48 |
| 0.0073469 | -666.078 |
| 0.00704544 | -618.281 |
| 0.0100932 | -820.841 |
| 0.0163214 | -954.573 |
| 0.0108302 | -829.445 |
| 0.00399056 | -376.939 |
| 0.0128259 | -762.235 |
| 0.0119994 | -838.744 |
| 0.0148876 | -1386.02 |
| 0.00866074 | -490.01 |
| 0.0105177 | -944.978 |
| 0.0117589 | -1056.33 |
| 0.00834461 | -1023.9 |
| 0.00741441 | -676.785 |
| 0.0107567 | -505.259 |
| 0.0179024 | -1598.48 |
| 0.00614998 | -350.423 |
| 0.0101639 | -434.583 |
| 0.0134426 | -787.764 |
| 0.00787509 | -808.193 |
| 0.00733028 | -692.803 |
| 0.0165941 | -1088.55 |
| 0.0200866 | -1418.25 |
| 0.0220854 | -1277.87 |
| 0.0172468 | -1181.28 |
| 0.00856186 | -928.005 |
| 0.0172025 | -1221.26 |
| 0.00393239 | -97.0184 |
| 0.0111311 | -941.049 |
| 0.0111603 | -917.427 |
| 0.0086315 | -497.488 |
| 0.00910985 | -748.641 |
| 0.0114016 | -603.33 |
| 0.00920404 | -570.291 |
| 0.00849422 | -921.667 |
| 0.0150693 | -899.99 |
| 0.011784 | -1211.27 |
| 0.0107878 | -1030.28 |
| 0.00571091 | -362.877 |
| 0.0176247 | -1435.58 |
| 0.00939337 | -511.629 |
| 0.0196564 | -902.346 |
| 0.0101283 | -577.304 |
| 0.000775268 | 40.3629 |
| 0.0158968 | -746.883 |
| 0.0170142 | -1669.2 |
| 0.00735864 | -449.639 |
| 0.00873973 | -433.439 |
| 0.00891854 | -589.266 |
| 0.0132341 | -1362.2 |
| 0.00744218 | -484.274 |
| 0.0117858 | -933.041 |
| 0.00745042 | -464.626 |
| 0.0136361 | -1076.23 |
| 0.00830485 | -391.223 |
| 0.00981928 | -723.393 |
| 0.00978643 | -752.025 |
| 0.00784566 | -584.928 |
| 0.0121991 | -552.595 |
| 0.00834528 | -419.084 |
| 0.00855268 | -506.303 |
| 0.00699681 | -764.75 |
| 0.0142109 | -861.961 |
| 0.0103891 | -635.359 |
| 0.0140022 | -862.542 |
| 0.0114624 | -884.217 |
| 0.0144058 | -887.342 |
| 0.00842825 | -650.781 |
| 0.0123699 | -1281.52 |
| 0.0117562 | -781.879 |
| 0.0108645 | -998.95 |
| 0.0142818 | -791.149 |
| 0.0164764 | -1378.92 |
| 0.0217088 | -1508.25 |
| 0.00900029 | -769.502 |
| 0.0105816 | -491.961 |
| 0.00928373 | -727.29 |
| 0.0103753 | -703.178 |
| 0.00860803 | -641.601 |
| 0.00932391 | -579.748 |
| 0.016343 | -946.001 |
| 0.0144063 | -1001.05 |
| 0.00843561 | -501.827 |
| 0.00806139 | -701.949 |
| 0.0128712 | -695.57 |
| 0.00956474 | -685.967 |
| 0.00868936 | -568.882 |
| 0.010996 | -788.565 |
| 0.0145881 | -858.799 |

##### **Figure S9. Cost-effectiveness acceptability curve of National PreCePT Programme**

The curve shows the probability of National PreCePT Programme being cost-effective at different cost-effectiveness threshold values.

COMBINED

Probability that the National PReCePT Programme is cost-effective

| Cost-effectiveness threshold, £ | pce |
| --- | --- |
| 0 | 0.9433 |
| 1000 | 0.9449 |
| 2000 | 0.9466 |
| 3000 | 0.9487 |
| 4000 | 0.951 |
| 5000 | 0.9529 |
| 6000 | 0.9536 |
| 7000 | 0.9543 |
| 8000 | 0.9556 |
| 9000 | 0.9568 |
| 10000 | 0.9579 |
| 11000 | 0.959 |
| 12000 | 0.96 |
| 13000 | 0.9609 |
| 14000 | 0.9617 |
| 15000 | 0.963 |
| 16000 | 0.9646 |
| 17000 | 0.9654 |
| 18000 | 0.9664 |
| 19000 | 0.9671 |
| 20000 | 0.9682 |
| 21000 | 0.9693 |
| 22000 | 0.9705 |
| 23000 | 0.9716 |
| 24000 | 0.972 |
| 25000 | 0.9727 |
| 26000 | 0.9731 |
| 27000 | 0.9735 |
| 28000 | 0.974 |
| 29000 | 0.9748 |
| 30000 | 0.9755 |
| 31000 | 0.9767 |
| 32000 | 0.9774 |
| 33000 | 0.9777 |
| 34000 | 0.9785 |
| 35000 | 0.979 |
| 36000 | 0.9797 |
| 37000 | 0.9804 |
| 38000 | 0.9809 |
| 39000 | 0.9812 |
| 40000 | 0.9815 |
| 41000 | 0.9822 |
| 42000 | 0.9825 |
| 43000 | 0.9829 |
| 44000 | 0.9831 |
| 45000 | 0.9835 |
| 46000 | 0.9839 |
| 47000 | 0.9843 |
| 48000 | 0.9845 |
| 49000 | 0.9846 |
| 50000 | 0.9851 |
| 51000 | 0.9856 |
| 52000 | 0.986 |
| 53000 | 0.9865 |
| 54000 | 0.9868 |
| 55000 | 0.9872 |
| 56000 | 0.9874 |
| 57000 | 0.9875 |
| 58000 | 0.9878 |
| 59000 | 0.988 |
| 60000 | 0.9883 |
| 61000 | 0.9887 |
| 62000 | 0.989 |
| 63000 | 0.9896 |
| 64000 | 0.9898 |
| 65000 | 0.9898 |
| 66000 | 0.99 |
| 67000 | 0.9903 |
| 68000 | 0.9907 |
| 69000 | 0.9907 |
| 70000 | 0.9907 |
| 71000 | 0.9908 |
| 72000 | 0.9913 |
| 73000 | 0.9917 |
| 74000 | 0.9918 |
| 75000 | 0.9918 |
| 76000 | 0.992 |
| 77000 | 0.9922 |
| 78000 | 0.9923 |
| 79000 | 0.9924 |
| 80000 | 0.9926 |
| 81000 | 0.9926 |
| 82000 | 0.9929 |
| 83000 | 0.993 |
| 84000 | 0.9932 |
| 85000 | 0.9933 |
| 86000 | 0.9934 |
| 87000 | 0.9935 |
| 88000 | 0.9936 |
| 89000 | 0.9936 |
| 90000 | 0.9937 |
| 91000 | 0.9937 |
| 92000 | 0.9938 |
| 93000 | 0.9941 |
| 94000 | 0.9942 |
| 95000 | 0.9942 |
| 96000 | 0.9944 |
| 97000 | 0.9944 |
| 98000 | 0.9944 |
| 99000 | 0.9945 |
| 100000 | 0.9945 |

##### **Figure S10. Cost-effectiveness acceptability curve of National PreCePT Programme for SCU/HDU units**

The curve shows the probability of National PreCePT Programme being cost-effective at different cost-effectiveness threshold values for SCU/HDU units.

SCU/HDU

Probability that the National PReCePT Programme is cost-effective

| Cost-effectiveness threshold, £ | pce |
| --- | --- |
| 0 | 0.7829 |
| 1000 | 0.788 |
| 2000 | 0.7939 |
| 3000 | 0.7981 |
| 4000 | 0.8027 |
| 5000 | 0.8067 |
| 6000 | 0.812 |
| 7000 | 0.8159 |
| 8000 | 0.8203 |
| 9000 | 0.8241 |
| 10000 | 0.8286 |
| 11000 | 0.8331 |
| 12000 | 0.8371 |
| 13000 | 0.8402 |
| 14000 | 0.8432 |
| 15000 | 0.8452 |
| 16000 | 0.8488 |
| 17000 | 0.8521 |
| 18000 | 0.8548 |
| 19000 | 0.8581 |
| 20000 | 0.861 |
| 21000 | 0.8639 |
| 22000 | 0.8671 |
| 23000 | 0.8693 |
| 24000 | 0.8714 |
| 25000 | 0.8739 |
| 26000 | 0.8764 |
| 27000 | 0.8789 |
| 28000 | 0.8816 |
| 29000 | 0.8841 |
| 30000 | 0.886 |
| 31000 | 0.8883 |
| 32000 | 0.8909 |
| 33000 | 0.8924 |
| 34000 | 0.8938 |
| 35000 | 0.8961 |
| 36000 | 0.8985 |
| 37000 | 0.9003 |
| 38000 | 0.9023 |
| 39000 | 0.9042 |
| 40000 | 0.9058 |
| 41000 | 0.9078 |
| 42000 | 0.9091 |
| 43000 | 0.9108 |
| 44000 | 0.9129 |
| 45000 | 0.9149 |
| 46000 | 0.9166 |
| 47000 | 0.9184 |
| 48000 | 0.9194 |
| 49000 | 0.9209 |
| 50000 | 0.922 |
| 51000 | 0.9235 |
| 52000 | 0.9247 |
| 53000 | 0.9257 |
| 54000 | 0.9267 |
| 55000 | 0.9278 |
| 56000 | 0.9296 |
| 57000 | 0.9309 |
| 58000 | 0.9324 |
| 59000 | 0.9333 |
| 60000 | 0.9345 |
| 61000 | 0.9354 |
| 62000 | 0.9367 |
| 63000 | 0.9374 |
| 64000 | 0.9384 |
| 65000 | 0.9386 |
| 66000 | 0.9398 |
| 67000 | 0.9412 |
| 68000 | 0.942 |
| 69000 | 0.9428 |
| 70000 | 0.9435 |
| 71000 | 0.9438 |
| 72000 | 0.9448 |
| 73000 | 0.9459 |
| 74000 | 0.9469 |
| 75000 | 0.9481 |
| 76000 | 0.9488 |
| 77000 | 0.9493 |
| 78000 | 0.95 |
| 79000 | 0.9511 |
| 80000 | 0.9514 |
| 81000 | 0.9521 |
| 82000 | 0.9529 |
| 83000 | 0.9535 |
| 84000 | 0.9541 |
| 85000 | 0.9551 |
| 86000 | 0.9555 |
| 87000 | 0.9564 |
| 88000 | 0.9569 |
| 89000 | 0.9576 |
| 90000 | 0.9583 |
| 91000 | 0.9592 |
| 92000 | 0.9597 |
| 93000 | 0.9604 |
| 94000 | 0.961 |
| 95000 | 0.9618 |
| 96000 | 0.9622 |
| 97000 | 0.9633 |
| 98000 | 0.9637 |
| 99000 | 0.964 |
| 100000 | 0.9645 |

##### **Figure S11. Cost-effectiveness acceptability curve of National PreCePT Programme for NICU units**

The curve shows the probability of National PreCePT Programme being cost-effective at different cost-effectiveness threshold values NICU units.

NICU

Probability that the National PReCePT Programme is cost-effective

| Cost-effectiveness threshold, £ | pce |
| --- | --- |
| 0 | 0.9952 |
| 1000 | 0.9952 |
| 2000 | 0.9953 |
| 3000 | 0.9954 |
| 4000 | 0.9954 |
| 5000 | 0.9954 |
| 6000 | 0.9956 |
| 7000 | 0.9956 |
| 8000 | 0.9956 |
| 9000 | 0.9957 |
| 10000 | 0.9957 |
| 11000 | 0.9958 |
| 12000 | 0.9958 |
| 13000 | 0.9959 |
| 14000 | 0.996 |
| 15000 | 0.996 |
| 16000 | 0.996 |
| 17000 | 0.996 |
| 18000 | 0.996 |
| 19000 | 0.9961 |
| 20000 | 0.9961 |
| 21000 | 0.9962 |
| 22000 | 0.9962 |
| 23000 | 0.9963 |
| 24000 | 0.9964 |
| 25000 | 0.9964 |
| 26000 | 0.9964 |
| 27000 | 0.9964 |
| 28000 | 0.9964 |
| 29000 | 0.9966 |
| 30000 | 0.9966 |
| 31000 | 0.9967 |
| 32000 | 0.9968 |
| 33000 | 0.9968 |
| 34000 | 0.9968 |
| 35000 | 0.9968 |
| 36000 | 0.9968 |
| 37000 | 0.9968 |
| 38000 | 0.9968 |
| 39000 | 0.9968 |
| 40000 | 0.9968 |
| 41000 | 0.9968 |
| 42000 | 0.9968 |
| 43000 | 0.9968 |
| 44000 | 0.9968 |
| 45000 | 0.9968 |
| 46000 | 0.9968 |
| 47000 | 0.9968 |
| 48000 | 0.9969 |
| 49000 | 0.9969 |
| 50000 | 0.9969 |
| 51000 | 0.9969 |
| 52000 | 0.9969 |
| 53000 | 0.997 |
| 54000 | 0.997 |
| 55000 | 0.997 |
| 56000 | 0.997 |
| 57000 | 0.997 |
| 58000 | 0.997 |
| 59000 | 0.997 |
| 60000 | 0.997 |
| 61000 | 0.9971 |
| 62000 | 0.9972 |
| 63000 | 0.9972 |
| 64000 | 0.9972 |
| 65000 | 0.9972 |
| 66000 | 0.9973 |
| 67000 | 0.9974 |
| 68000 | 0.9974 |
| 69000 | 0.9974 |
| 70000 | 0.9974 |
| 71000 | 0.9974 |
| 72000 | 0.9974 |
| 73000 | 0.9975 |
| 74000 | 0.9975 |
| 75000 | 0.9975 |
| 76000 | 0.9975 |
| 77000 | 0.9975 |
| 78000 | 0.9975 |
| 79000 | 0.9976 |
| 80000 | 0.9976 |
| 81000 | 0.9976 |
| 82000 | 0.9976 |
| 83000 | 0.9976 |
| 84000 | 0.9976 |
| 85000 | 0.9976 |
| 86000 | 0.9976 |
| 87000 | 0.9977 |
| 88000 | 0.9977 |
| 89000 | 0.9977 |
| 90000 | 0.9977 |
| 91000 | 0.9977 |
| 92000 | 0.9977 |
| 93000 | 0.9977 |
| 94000 | 0.9977 |
| 95000 | 0.9977 |
| 96000 | 0.9977 |
| 97000 | 0.9978 |
| 98000 | 0.9978 |
| 99000 | 0.9978 |
| 100000 | 0.9978 |

1.	Bickford CD, Magee LA, Mitton C, et al. Magnesium sulphate for fetal neuroprotection: a cost-effectiveness analysis. *BMC Health Services Research* 2013; **13**: 527.
